# Mutation hotspots, geographical and temporal distribution of SARS-CoV-2 lineages in Brazil, February 2020-2021: insights and limitations from uneven sequencing efforts

**DOI:** 10.1101/2021.03.08.21253152

**Authors:** Vinícius Bonetti Franceschi, Patrícia Aline Gröhs Ferrareze, Ricardo Ariel Zimerman, Gabriela Bettella Cybis, Claudia Elizabeth Thompson

## Abstract

The COVID-19 pandemic has already reached approximately 110 million people and it is associated with 2.5 million deaths worldwide. Brazil is the third worst-hit country, with approximately 10.2 million cases and 250 thousand deaths. International efforts have been established to share information about SARS-CoV-2 epidemiology and evolution. However, sequencing facilities and research investments are very heterogeneous across different regions and countries. The understanding of the SARS-CoV-2 evolution plays a significant role in the development of effective strategies for public health and disease management. We aimed to analyze the available and high-quality genome sequences from Brazil between February 2020 and February 2021 to identify mutation hotspots, geographical and temporal distribution of SARS-CoV-2 lineages by using phylogenetics and phylodynamics analyses. We describe heterogeneous and episodic sequencing efforts, the progression of the different lineages along time, evaluating mutational spectra and frequency oscillations derived from the prevalence of novel and specific lineages across different Brazilian regions. We found at least seven major (1-7) and two minor clades (4.2 and 5.3) related to the six most prevalent lineages in the country and described its spatial distribution and dynamics. The emergence and recent frequency shift of lineages (P.1 and P.2) containing mutations of concern in the spike protein (*e. g*., E484K, N501Y) draws attention due to their association with immune evasion and enhanced receptor binding affinity. Improvements in genomic surveillance are of paramount importance and should be extended in Brazil to better inform policy makers and enable evidence-based decisions to fight the COVID-19 pandemic.

## Introduction

After its initial emergence in China in late 2019 ^1^, *Severe Acute Respiratory Syndrome 2 Virus* (SARS-CoV-2) has spread rapidly around the world causing the COVID-19 pandemic ^2^. New epicenters of the disease have been established throughout 2020, mainly in Europe, USA, and South America ^3,4^. As of February 22, 2021, more than 110 million cases and approximately 2.5 million deaths worldwide have been confirmed ^5^. Several countries are currently experiencing second waves of infections, decreasing optimism regarding a brief solution to the pandemic.

Since the sequencing of the first SARS-CoV-2 genome ^6^, international efforts have been established through data sharing in the GISAID database ^7^. These sequencing and metadata information were made public and have enabled the study of the viral spread pattern through space and time. However, sequencing facilities and research investments are very heterogeneous across the world. Asian, European, North American and Oceanian countries have contributed with more data proportionally to the number of cases ^8^, while African and South American genomic surveillance have been more limited. Disparities are even deeper on an individual country basis. For instance, while the United Kingdom leads sequencing efforts (5.85% of cases sequenced), Brazil has sequenced only 0.03% of all its cases, despite being the third worst-hit country, with approximately 10.2 million cases and 250 thousand deaths.

Many studies were performed to characterize early viral introductions and transmission dynamics in several countries (*e. g*. China ^9^, USA ^10–12^, Australia ^13^, Italy ^14^, United Kingdom ^15,16^). In Brazil, SARS-CoV-2 arrived officially on February 25, 2020, in a returning traveller from Italy, and early efforts were made both at the national ^17^ and regional levels ^18,19^ to further characterize viral introduction and spread. B.1 and derived lineages were prevalent in the country at the beginning of the pandemic and significant movements between state borders after international travel restrictions have been demonstrated ^17^. Unfortunately, little is known about the viral evolution in the entire Brazilian territory after these earliest studies.

More recently, new SARS-CoV-2 lineages have emerged and are considered as “Variants Of Concern” (VOC), mainly those carrying mutations in the spike (S) glycoprotein due to its role in binding to the human ACE2 receptor (hACE2). Up to February 2021, there are three VOCs described worldwide, namely: B.1.1.7, B.1.351, and P.1. The former (B.1.1.7) emerged in England in mid-September 2020 and it is characterized by 14 lineage-specific amino acid substitutions, especially N501Y (a key contact residue interacting with hACE2) and P681H (one of four amino acids comprising the insertion that creates a novel furin cleavage site between S1 and S2) ^20^. The second (B.1.351) emerged in South Africa in October 2020 and harbor a constellation of mutations in the Receptor Binding Domain (RBD) (especially K417N, E484K and N501Y) ^21^. The more recent lineage is P.1, derived from B.1.1.28, a widespread lineage from Brazil. It was recently reported in returning travelers from Manaus (Amazonas, Brazil), after arriving in Japan. It has the same three mutations (except for K417T instead of K417N) in the RBD as B.1.351, but it arose independently ^22^. Importantly, B.1.351 and P.1 carry the E484K mutation associated with escape from neutralizing antibodies ^23–25^. Recently, a E484K harboring virus was identified in a reinfected patient ^26^ from Brazil, confirming the ability to evade naturally developed antibodies from previous infection as well. Moreover, all three VOC lineages harbor N501Y mutation, already associated with enhanced receptor binding affinity ^27^, which could lead to increased infectiousness.

It is believed that after more than a year of its emergence, some mutations of SARS-CoV-2 (*e. g*. E484K and N501Y) have been positively selected, since they may confer adaptive advantages leading to convergent evolution in different lineages spreading across multiple countries ^28^. Despite the low sequencing rate, a deeper analysis of mutations and lineages throughout the Brazilian states would allow a better understanding of viral diversity and spread patterns inside the country. Thus, we aimed to identify mutation hotspots, geographical and temporal distribution of SARS-CoV-2 lineages in the Brazilian territory by using phylogenetics and phylodynamics analyses from high-quality SARS-CoV-2 genome sequences.

## Material and methods

### SARS-CoV-2 genomes and epidemiological data retrieval

Complete SARS-CoV-2 genomes (>29,000 bp) and the associated metadata were obtained from the GISAID database. Considering 2,751 available sequences from Brazil submitted until February 16 2021, 2732 were retrieved applying filters for human host and complete collection date. Number of cases per state per day and across Brazil were downloaded from https://covid19br.wcota.me/en/ ^29^ on the same date. This initiative aggregates data from the Brazilian Ministry of Health and epidemiological bulletins of each federative unit.

### Mutation analysis

The GenBank RefSeq sequence NC_045512.2 from Wuhan (China) was used as the reference for our analysis. Single nucleotide polymorphisms (SNPs) and insertions/deletions (INDELs) were assessed by using snippy variant calling pipeline v4.6.0 (https://github.com/tseemann/snippy), which uses FreeBayes v1.3.2 ^30^ as variant caller and snpEff v5.0 ^31^ to annotate and predict the effects of variants on genes and proteins. Mutations and lineages were concatenated with associated metadata and counted by Brazilian states using custom Python and R scripts. Histogram of SNPs were generated after running MAFFT v7.471 ^32^ alignment using msastats.py script, and plotAlignment and plotSNPHist functions (https://github.com/laduplessis/SARS-CoV-2_Guangdong_genomic_epidemiology/).

### Phylogenetics analysis

All available SARS-CoV-2 genomes (537,360 sequences) were retrieved from GISAID on February 16, 2021. These sequences were then subjected to analysis inside NextStrain ncov pipeline (https://github.com/nextstrain/ncov) ^33^ through a Brazilian-focused subsampling scheme using time- and worldwide-representative contextual samples.

In this workflow, sequences were filtered out based on high divergence, incompleteness and sampling date availability. Next, filtered genomes were aligned using nextalign v0.1.6 (https://github.com/neherlab/nextalign) and their ends were masked (100 positions in the beginning, 50 in the end). Maximum likelihood (ML) phylogenetics tree was built using IQ-TREE v2.0.3 ^34^, employing the best-fit model of nucleotide substitution as selected by ModelFinder ^35^. The root of the tree was placed between lineage A and B (Wuhan/WH01/2019 and Wuhan/Hu-1/2019). The clock-like behavior of the inferred tree was inspected using TempEst v1.5.3 ^36^ to generate the root-to-tip regression against sampling dates (correlation coefficient = 0.83, R^2^ = 0.68). Sequences that deviate more than four interquartile ranges from the root-to-tip regression were removed from the analysis. The subsampled time-scaled ML phylogenetics tree was generated using TreeTime v0.8.1 under a strict clock and a skyline coalescent prior with a rate of 8×10^−4^ substitutions per site per year ^37^. Results were then exported to JSON format to enable interactive genetic and geographical visualization using Auspice. Additionally, ML and time-stamped trees were visualized using FigTree v1.4 (http://tree.bio.ed.ac.uk/software/figtree/) and ggtree R package v2.0.4 ^38^. We identified global lineages using the dynamic nomenclature ^39^ implemented in Pangolin v2.2.2 (https://github.com/cov-lineages/pangolin).

### Bayesian phylogeographic and phylodynamic analysis

All major clades associated with massive community transmission in Brazil (clades 3-6) as defined by the previous maximum likelihood analysis were included in this analysis. Clade 7 was discarded due to the low correlation of genetic distances and sampling dates (R^2^ = 2.11×10^−2^, correlation coefficient = 0.1488). Additionally, most of its sequences ranged from 2020.8 and 2021.1 and had high divergence, suggesting important undersampling for this clade (represented by sequences from P.2 lineage). The spatiotemporal diffusion of these important circulating lineages through Brazil were separately estimated for each clade using a Bayesian Markov Chain Monte Carlo (MCMC) approach as implemented in BEAST v1.10.4 ^40^, using the BEAGLE library v3 ^41^ to enhance computational time. Time-scaled Bayesian trees were estimated in BEAST using: HKY+_Γ_ nucleotide substitution model ^42^, a strict molecular clock model with a Continuous Time Markov Chain (CTMC) ^43^ prior (mean rate = 8×10^−4^), and a parametric Exponential Growth coalescent tree prior ^44^. Although it is not the most appropriate model to reconstruct the demographic and evolutionary processes, it was used given the difficulty of convergence in more complex models due to the lack of sufficient evolutionary information contained in the data. For clade 4, we used a non-parametric Bayesian Skygrid prior with 10 parameters ^45^.

Viral migrations across time were reconstructed using a Brownian random walk continuous phylogeographic model ^46,47^ to generate a posterior distribution of 1,000 trees whose internal nodes are associated with geographic coordinates. We assigned sampling coordinates using the centroid point of each municipality (when available) and the centroid of the state’s capital (when the municipality was unavailable) using a random jitter window size of 0.01 to add a small amount of noise to duplicated sampling coordinates assigned to the tips of the tree.

Two MCMC chains were run for >100 million generations and convergence of the MCMC chains was inspected using Tracer v1.7.1 ^48^. After removal of 10% burn-in, log and tree files were combined using LogCombiner v1.10.4 ^40^. Maximum clade credibility (MCC) trees were generated using TreeAnnotator v1.10.4 ^40^. The seraphim R package ^49^ and SPREAD3 ^50^ were used to extract and map the spatiotemporal information embedded in MCC trees. Finally, these trees were visualized using the ggtree R package v2.0.4 ^38^.

## Results

### Distribution of Brazilian sequences through time and space

Sequencing efforts from Brazil were concentrated mainly in the first epidemic wave (March to April, 2020) (Figure 1A and 1B). In March, 503 genomes (8.64% of the confirmed cases) and in April, 942 sequences (1.16% of the cases) were sampled. All following months fell below 1% of sequencing rate (Table S1). All Brazilian states sequenced less than 0.1% of the confirmed cases through the first year of the pandemic (Figure 1C). From the Southeast region, the states of Rio de Janeiro (0.09%) and São Paulo (0.06%) have led the country’s sequencing initiatives, followed by Rio Grande do Sul (0.045%) from the South region, Amazonas (0.041%) from the North region and Pernambuco (0.040%) from the Northeast region. The Centre-West was the region with the lowest sequencing rate (Figure 1C, Table S2).

**Figure 1.**
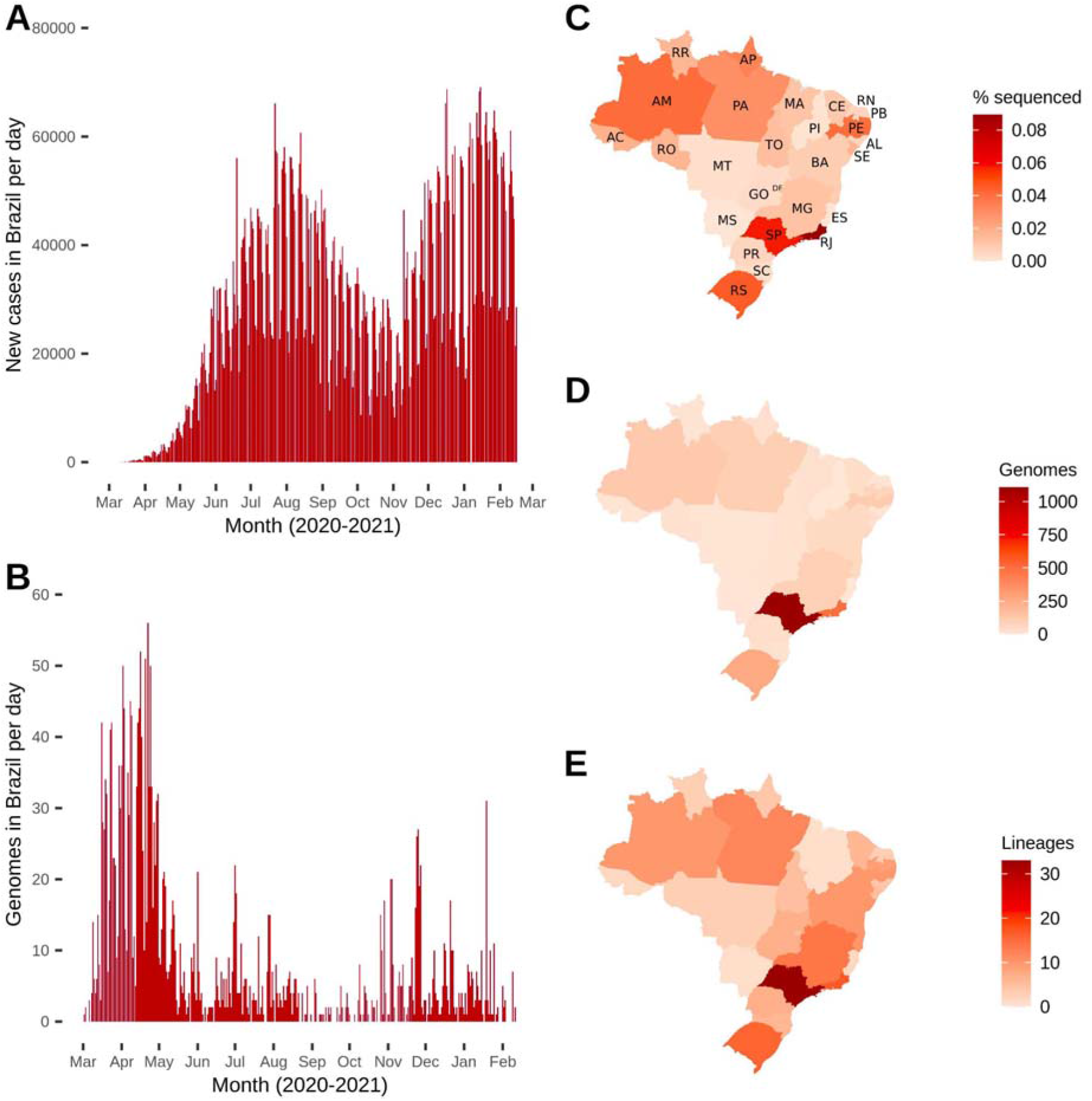
Distribution of Brazilian genomes through time (end-February 2020 to mid-February 2021) and space (Brazilian states). (A) Number of new cases per day in Brazil over time. (B) Number of genomes (sequenced cases) in Brazil over time (date of sample collection). (C) Fraction of genomes sequenced related to number of cases per Brazilian state. (D) Total number of genomes deposited per Brazilian state. (E) Total number of different SARS-CoV-2 lineages detected per Brazilian state. State abbreviations: AC=Acre; AL=Alagoas; AM=Amazonas; Amapá=AP; BA=Bahia; CE=Ceará; DF=Distrito Federal; ES=Espírito Santo; GO=Goiás; MA=Maranhão; MG=Minas Gerais; MS=Mato Grosso do Sul; MT=Mato Grosso; PA=Pará; PE=Pernambuco; PB=Paraíba; PI=Piauí; PR=Paraná; RJ=Rio de Janeiro; RN=Rio Grande do Norte; RO=Rondônia; RR=Roraima; RS=Rio Grande do Sul; SC=Santa Catarina; SE=Sergipe; SP=São Paulo; TO=Tocantins.

The Southeast region contributed with 1,704 genomes (62.53%), followed by Northeast (n=359; 13.17%), South (n=319; 11.71%), North (n=310; 11.38%), and Centre-West (1.21%) regions (Figure 1D, Table S3). In total, 59 different lineages were detected in Brazil, and the states sequencing more genomes (São Paulo, Rio de Janeiro and Rio Grande do Sul) detected higher numbers of circulating lineages (33, 17 and 16, respectively) (Figure 1E, Table S3). Importantly, all States that sequenced at least 100 genomes identified _≥_ 10 lineages (Table S3).

### High frequency mutations

A total of 3,919 mutations were detected across the 2,731 Brazilian genomes and only 354 (12.96%) occurred in >5 sequences, 44 (1.61%) in >50 genomes, and 38 (1.39%) in >100 sequences (Figure 2, Table 1). Twenty-five (65.79%) of these 38 mutations were non-synonymous. Of these, 11 (44.0%) were in the spike protein, 5 (20.0%) in the nucleocapsid protein, and 5 (20.0%) in the ORF1ab polyprotein (Table 1).

**Table 1.** Mutations of Brazilian genomes found in > 100 genomes, associated effects on encoded proteins and major associated lineages.

| REF | Genomic position | ALT | Effect | Amino acid | Gene | Product | Number of sequences | % in Brazilian genomes | Major associated lineages |
| --- | --- | --- | --- | --- | --- | --- | --- | --- | --- |
| A | 23403 | G | Missense | D614G | S | Surface glycoprotein | 2651 | 97.07 | B.1 and derived |
| C | 14408 | T | Synonymous | L4715L | ORF1ab | RdRp | 2648 | 96.96 | B.1 and derived |
| C | 3037 | T | Synonymous | F924F | ORF1ab | nsp3 | 2635 | 96.48 | B.1 and derived |
| C | 241 | T | Intergenic | - | - | - | 2374 | 86.93 | B.1 and derived |
| GGG | 28881 | AAC | Missense | RG203-204KR | N | N phosphoprotein | 2326 | 85.17 | B.1.1 and derived |
| G | 25088 | T | Missense | V1176F | S | Surface glycoprotein | 1299 | 47.56 | B.1.1.28, B.1.1.143, B.1.1.94, P.1, and P.2 |
| T | 29148 | C | Missense | I292T | N | N phosphoprotein | 888 | 32.52 | B.1.1.33, B.1.1.314, N.1, N.4 |
| T | 27299 | C | Missense | I33T | ORF6 | ORF6 protein | 885 | 32.41 | B.1.1.33, B.1.1.314 |
| C | 12053 | T | Missense | L3930F | ORF1ab | nsp7 | 761 | 27.87 | B.1.1.28, B.1.1.74, B.1.1.143, P.2 |
| G | 23012 | A | Missense | E484K | S | Surface glycoprotein | 312 | 11.42 | B.1.1.33, P.1, P.2 |
| T | 26149 | C | Missense | S253P | ORF3a | ORF3a protein | 249 | 9.12 | B.1.1.28, P.1 |
| A | 6319 | G | Synonymous | P2018P | ORF1ab | nsp3 | 244 | 8.93 | B.1.1.28, P.1 |
| C | 28253 | T | Synonymous | F120F | ORF8 | ORF8 protein | 222 | 8.13 | B.1.1.28, B.1.1.33, P.2 |
| G | 28975 | T | Missense | M234I | N | N phosphoprotein | 217 | 7.95 | B.1.1.28, P.2 |
| G | 28628 | T | Missense | A119S | N | N phosphoprotein | 211 | 7.73 | P.2 |
| C | 11824 | T | Synonymous | I3853I | ORF1ab | nsp6 | 207 | 7.58 | P.2 |
| T | 10667 | G | Missense | L3468V | ORF1ab | 3C-like proteinase | 206 | 7.54 | P.2 |
| A | 12964 | G | Synonymous | G4233G | ORF1ab | nsp9 | 176 | 6.44 | P.2 |
| A | 6613 | G | Synonymous | V2116V | ORF1ab | nsp3 | 137 | 5.02 | B.1.1.28, P.1 |
| GTCTGG<br>TTTT | 11287 | G | Deletion | 3675-3677<br>SGF | ORF1ab | nsp6 | 130 | 4.76 | B.1.1.7, P.1 |
| C | 29754 | T | Intergenic | - | - | - | 117 | 4.28 | P.2 |
| AGTAGG<br>G | 28877 | TCTA<br>AAC | Missense | RG203-204KR | N | N phosphoprotein | 116 | 4.25 | B.1.1.28, P.1 |
| C | 21614 | T | Missense | L18F | S | Surface glycoprotein | 116 | 4.25 | B.1.1.28, P.1 |
| G | 17259 | T | Missense | S5665I | ORF1ab | Helicase | 116 | 4.25 | P.1 |
| A | 23063 | T | Missense | N501Y | S | Surface glycoprotein | 115 | 4.21 | B.1.1.7, P.1 |
| C | 21638 | T | Missense | P26S | S | Surface glycoprotein | 113 | 4.14 | P.1 |
| T | 733 | C | Synonymous | D156D | ORF1ab | Leader protein | 111 | 4.06 | P.1 |
| C | 13860 | T | Missense | T4532I | ORF1ab | RdRp | 111 | 4.06 | P.1 |
| C | 24642 | T | Missense | T1027I | S | Surface glycoprotein | 111 | 4.06 | P.1 |
| C | 12778 | T | Synonymous | Y4171Y | ORF1ab | nsp9 | 111 | 4.06 | P.1 |
| C | 21621 | A | Missense | T20N | S | Surface glycoprotein | 111 | 4.06 | P.1 |
| C | 28512 | G | Missense | P80R | N | N phosphoprotein | 110 | 4.03 | P.1 |
| A | 5648 | C | Missense | K1795Q | ORF1ab | nsp3 | 109 | 3.99 | P.1 |
| C | 23525 | T | Missense | H655Y | S | Surface glycoprotein | 109 | 3.99 | P.1 |
| G | 28167 | A | Missense | E92K | ORF8 | ORF8 protein | 108 | 3.95 | P.1 |
| G | 21974 | T | Missense | D138Y | S | Surface glycoprotein | 107 | 3.92 | B.1.1.33, P.1 |
| G | 22132 | T | Missense | R190S | S | Surface glycoprotein | 105 | 3.84 | P.1 |
| C | 2749 | T | Synonymous | D828D | ORF1ab | nsp3 | 105 | 3.84 | P.1 |

REF: Reference nucleotide(s); ALT: Replaced nucleotide(s); UTR= Untranslated region; ORF=Open reading frame; S: Spike; N: Nucleocapsid; nsp: nonstructural protein; RdRp: RNA-dependent RNA polymerase.

**Figure 2.**
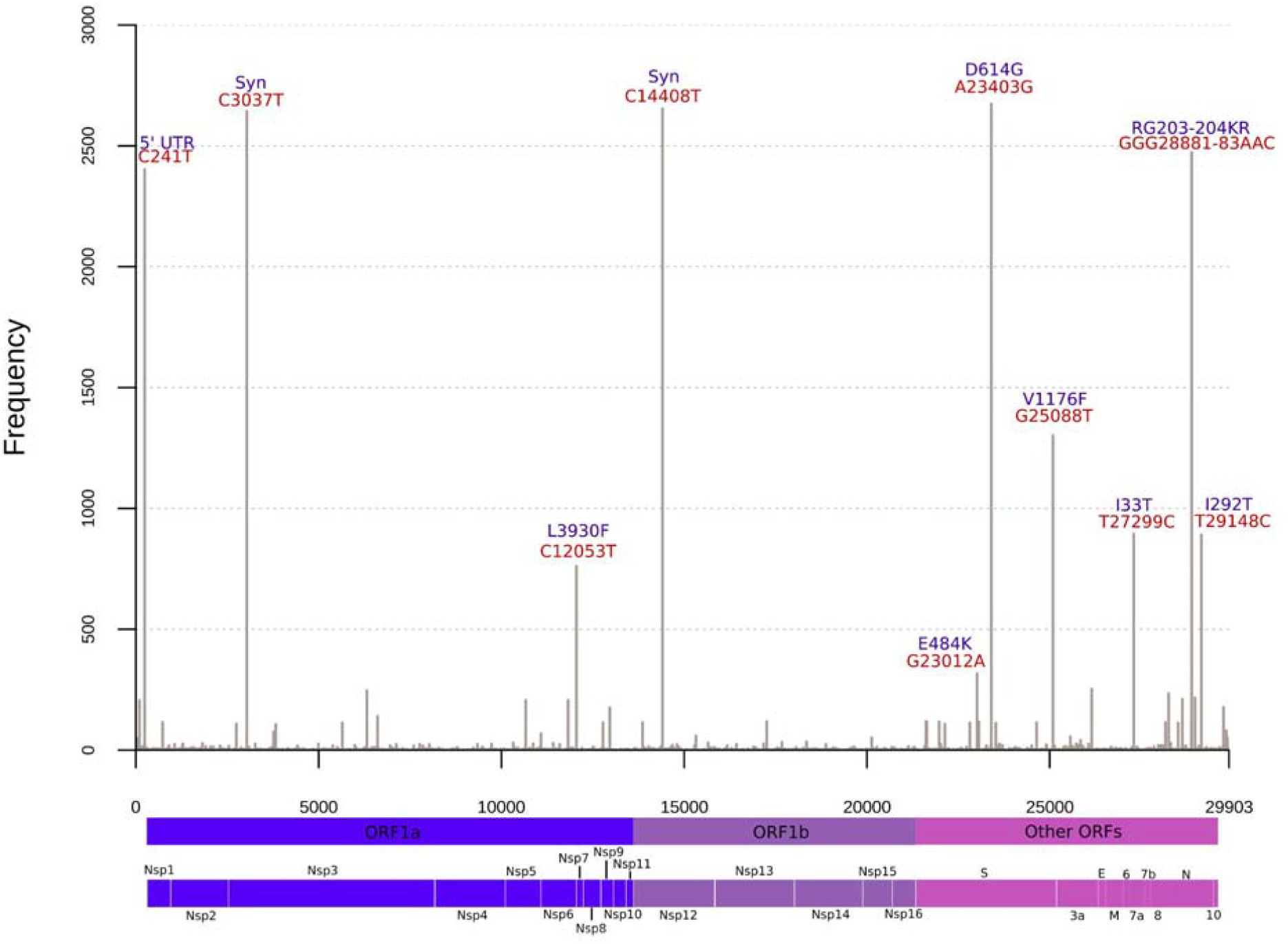
High frequent mutations across Brazilian sequences. Nucleotide replacements occurring in >250 genomes are indicated in red and associated amino acid substitutions in blue. Other mutations occurring in less than 250 genomes but more than 100 are indicated in Table 1. Syn: Synonymous.

Three mutations were found in >95% of the genomes: A23403G (S:D614G), C14408T (ORF1ab:L4715), and C3037T (ORF1ab:F924), which are signatures of the B.1 and derived lineages that spread early in the pandemic. The adjacent replacement GGG28881AAC (N:RG203-204KR) was found in 85.17%, representing a clear signature of B.1.1 lineage. The defining-mutations of the Brazilian most widespread lineages (B.1.1.28 and B.1.1.33) were also found in high abundance. The G25088T (S:V1176F) replacement from B.1.1.28 occurred in 47.56% of all sequences, while T29148C (N:I292T) and T27299C (ORF6:I33T) from B.1.1.33 in _≈_32.5%. The G23012A (S:E484K) mutation in the Receptor Binding Domain (RBD) of spike that recently emerged independently in three Brazilian lineages (B.1.1.33, P.1 and P.2) is already among the most frequent detected up to February, 2021 (11.42%). The E484K viruses have been spreading mostly between mid-2020 up to early-2021. Additionally, the multiple lineage-defining mutations found in emergent P.1 and P.2 lineages from Brazil were observed in >100 and >200 genomes, respectively (Figure 2, Table 1).

### Lineage dynamics of Brazilian epidemic

In March 2020, the majority of Brazilian sequences belonged to 3 lineages: B.1 (n=101; 20.08%), B.1.1.28 (n=156; 31.01%) and B.1.1.33 (n=131; 26.04%). The first was probably introduced in Brazil through multiple imports from other continents, and the others probably emerged from community transmission inside the country ^17^. Between April and August, >75% of all sequences per month were classified as B.1.1.28 or B.1.1.33. During October and November, the B.1.1.28 derived lineage P.2 was the most prevalent (n=32; 37.65% and n=92; 40.71%), while B.1.1.28 and B.1.1.33 represented together <50% of the genomes. From December 2020 onward, the VOC P.1 emerged in Manaus and has been established, together with P.2, as the most prevalent lineages represented by sequencing data (Figure 3A).

**Figure 3.**
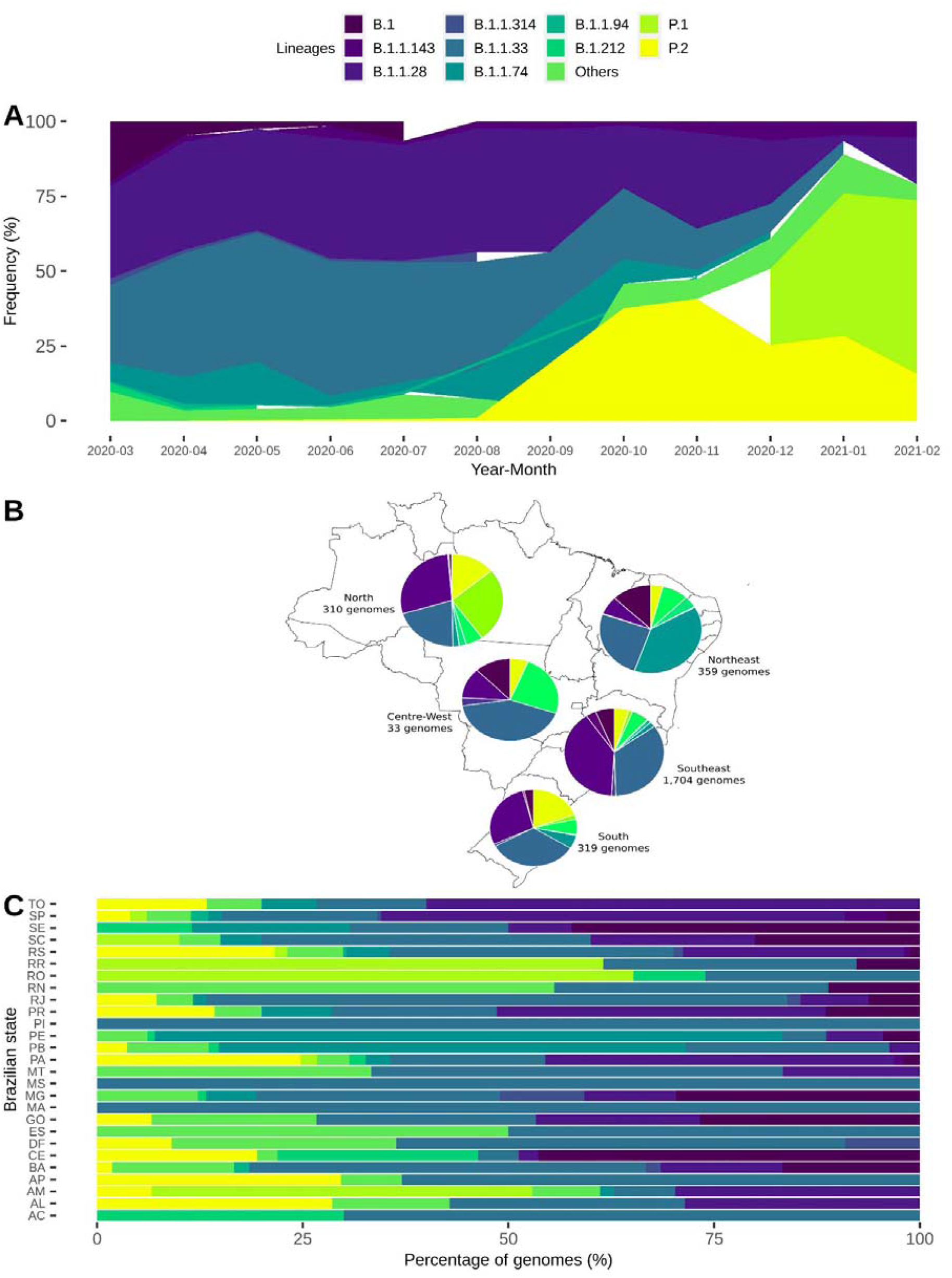
Distribution of the 10 most prevalent SARS-CoV-2 lineages (n>25) inside Brazil from end-February 2020 to mid-February 2021. (A) Frequency of these lineages through time in the entire Brazil. (B) Map showing the fraction of each of these lineages across all five Brazilian regions. (C) Distribution of these lineages across all Brazilian states, proportional to the number of sequenced genomes.

Regarding distribution between the five different Brazilian regions, the Southeast and Southern regions sequenced a larger proportion of B.1.1.28 and B.1.1.33 viruses. The Northeast apparently has a slightly different dynamics, since B.1.1.74 (n=138; 38.44%) is the most prevalent lineage followed by B.1.1.33 (n=91; 25.35%). In the Northern region, P.2 is already the second most prevalent lineage (n=81; 26.13%) (Figure 3B and 3C), but this may be related to the low quantity of sequences from the beginning of the pandemic, and the higher surveillance in the region at present, due to enhanced sequencing efforts after the emergence of P.1. In the Centre-West, the extremely low sequencing rate prevents us from making any assumptions about the genetic diversity of the circulating lineages (Figure 3B).

Statewide view shows the wide distribution of lineages B.1.1.28, B.1.1.33 and P.2 across almost all Brazilian states. Although the P.1 lineage has been represented by genomes from only 7 states (AM, PA, RS, RO, RR, SC, and SP) up to February 16 2021, other states have already reported its detection after this date. States reporting a higher proportion of B.1 lineage (CE, GO, MG, SC, and SE) have concentrated their sequencing efforts mainly in the early phase of the pandemic. B.1.1.74 is overrepresented (>50%) in two states from the Northeast (PE and PB), suggesting a more geographically restricted distribution. Additionally, the contribution of other lineages that are not among the 10 most frequent in the country are apparently limited (Figure 3C and Figure S1).

The time between the first and last detection of the 18 lineages represented by more than five Brazilian genomes varied markedly, with a mean of 186.28 days, median of 179.0 days and standard deviation of 119.58 days. Eleven of the 18 lineages were sampled in different states between the first and last detection, suggesting its spread through the Brazilian territory (Table S4). B and B.1 lineages have presented a relatively low time of spread, while several B.1.1 derived lineages probably have spreaded for a longer period (*e. g*, B.1.1.143, B.1.1.28, B.1.1.314, B.1.1.33, B.1.1.74, B.1.1.94) (Figure S2). B.1.1.7 (UK-associated lineage) was firstly detected in the country in mid-December ^51^, while P.1 was found in November in Manaus (Amazonas, Brazil) and Japan ^22^. P.2 was discovered in mid-April 2020, but has been increasing rapidly in frequency since August 2020 (Figure 3A, Figure S2).

### Phylogenetic analysis of SARS-CoV-2 genomes from Brazil

We obtained 10,573 time-, geographical- and genetic-representative genomes to proceed phylogenetic inferences. Of these, 1,135 were from Africa, 1,963 were from Asia, 3,248 from Europe, 612 from North America, 285 from Oceania, and 3,330 from South America. Among the latter, 2,350 were from Brazil.

The maximum likelihood tree showed the evolutionary diversity of SARS-CoV-2 sequences across Brazilian territory (Figure 4A). Almost all sequences from Brazil harbor the S:D614G and ORF1b:P314L, which were imported from other continents to Brazil (mainly to Southeastern states) in the early epidemic wave of COVID-19. After the importation of B.1.1 lineages characterized by N:R203K and N:G204R mutations, community transmission massively occurred and gave rise to B.1.1.28 (S:V1176F) and B.1.1.33 lineages (ORF6:I33T and N:I292T), widely distributed in Brazilian regions along the first year of the pandemic (Figure 4A and 4B). B.1.1.28 have diversified in two lineages, P.1 (20J/501Y.V3) and P.2, which are already widely represented by many sequences and distributed across all regions. The larger branch length leading to P.1 lineage draws attention and its emergence is probably driven by an accelerated molecular evolution (Figure 4B and 5A).

**Figure 4.**
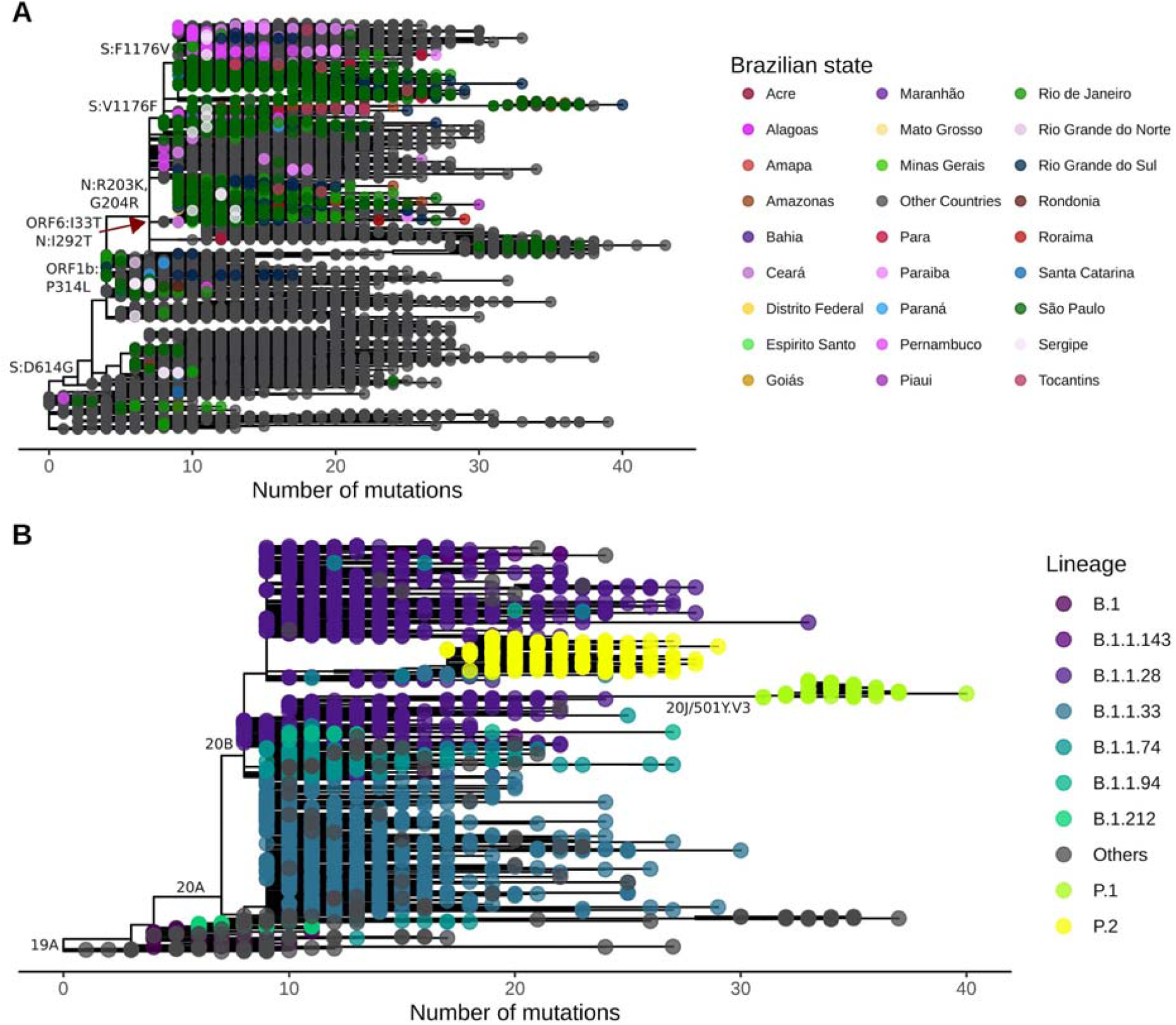
Evolutionary distribution of SARS-CoV-2 genomes from Brazil. (A) Maximum likelihood phylogenetics tree of 2,346 Brazilian sequences and 8,227 additional global representative genomes. States belonging to each specific Brazilian region are colored using similar colors (Centre-West: yellow; North: red; Northeast: purple and pink; South: blue; Southeast: green). Key mutations are represented in the respective branches. (B) Maximum likelihood phylogenetics tree dropping sequences from other countries and highlighting the most frequent Brazilian lineages presented in Figure 3. Nextstrain clades are represented in key branches. Evolutionary rate is represented by the number of mutations (divergences) related to SARS-CoV-2 reference sequence (NC_045512.2).

We estimated an evolutionary rate of 7.76×10^−4^ substitutions per site per year (23.22 mutations per year) for the Brazilian-focused subsampling (Figure 5A). Since the end of 2020, two evolutionary patterns are observed in the root-to-tip regression of sampling dates. Despite the majority of sequences following the expected substitution rate for SARS-CoV-2, we observe a rise in P.1 (20J/501Y.V3) and B.1.1.7 (20I/501Y.V1) abundance, both characterized by an abnormal clock rate (Figure 5A) and a constellation of mutations in the spike protein. Time-resolved maximum likelihood tree highlighted the importation of lineages in the early phase of the pandemic and its rapid diversification through community transmission inside the country, leading to the spread of two main lineages (B.1.1.28 and B.1.1.33) within and between state borders (Figure 5B) and a more restricted circulation of B.1.1.74 lineage in the Northeast. A time-resolved phylogeny considering only Brazilian sequences reinforces the nationwide distribution of multiple SARS-CoV-2 lineages and clades (Figure 5C and Figure S3).

**Figure 5.**
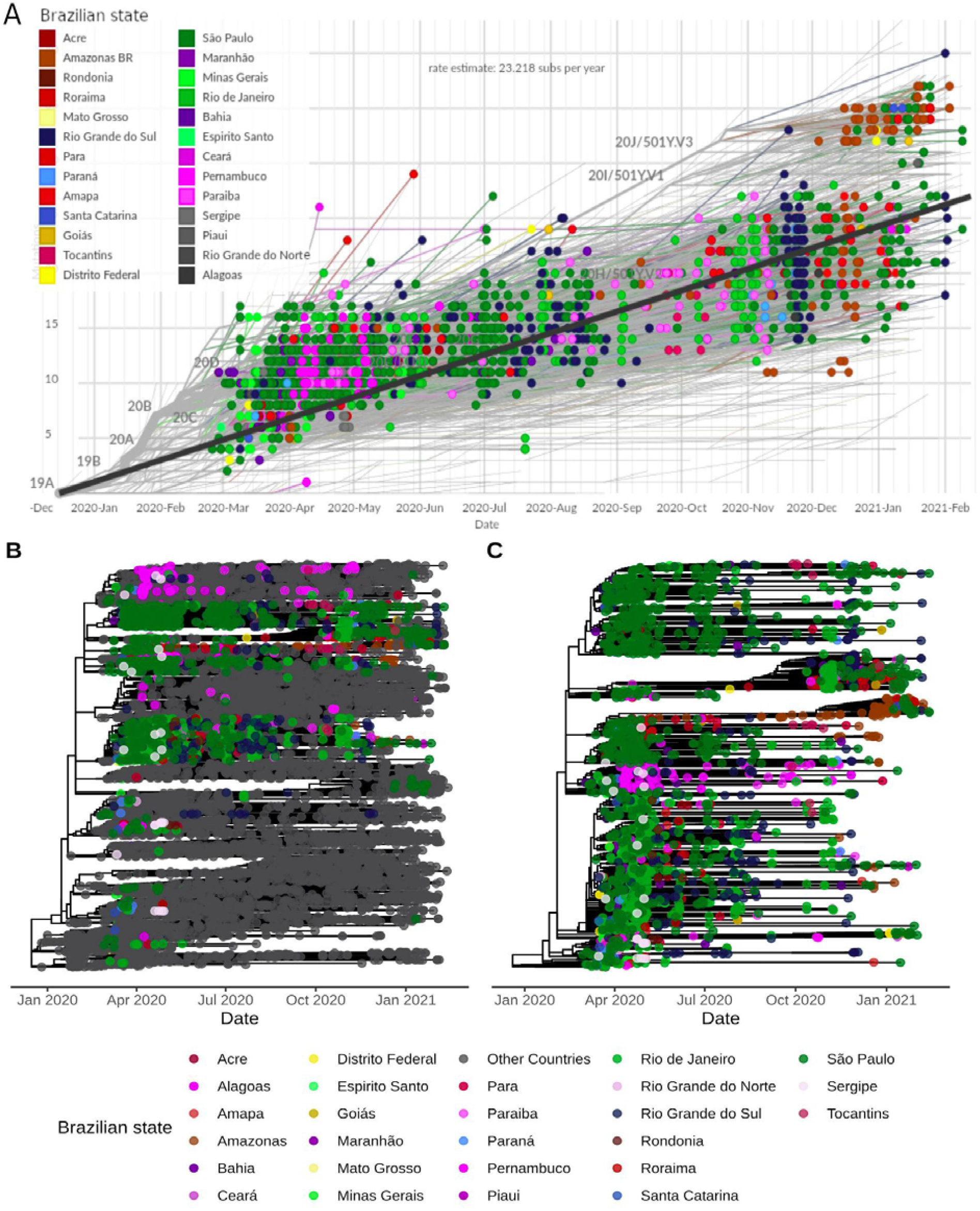
Molecular clock estimates of SARS-CoV-2 genomes from Brazil. (A) Root-to-tip regression of genetic distances (in number of mutations) against sampling dates filtered by Brazilian sequences. States belonging to each specific Brazilian region are colored using similar colors (Centre-West: yellow; North: red; Northeast: pink and gray; South: blue; Southeast: green). Nextstrain clades are represented in key branches. (B) Time-resolved Maximum Likelihood phylogenetics tree of the 10,573 genomes included colored by Brazilian states and other countries. (C) Maximum likelihood phylogenetics tree considering only the dynamics inside Brazil. In (B) and (C), state colorings follow the same scheme as (A), except for Northeast, where purple and pink colors define sequences from this region. Tree topology remains the same for these two trees, but node ordering is slightly different.

We found at least seven major clades (1-7) (Figure 6) and two minor (4.2 and 5.3) (Figure S4) related to the six most prevalent Brazilian lineages. Clade 1 (B.1 lineage) is represented by 33 genomes, >50% from the Southeast with a few introductions to other regions and a restricted time of spread until around May 2020 (Figure 6B). Clade 2 includes 98 genomes and is associated with and B.1.212 lineages. B.1 (Clade 2.1) sequences from this clade are mostly restricted to Southern and Southeast Brazil, while B.1.212 (Clade 2.2) has spread mainly in the Northeast and Northern regions (Figure 6C). Clade 3 is represented by 804 genomes, mostly from B.1.1.33 lineage. It reaches an unprecedented dissemination through Brazil since March, whose introductions in different regions and states were mostly driven by the Southeast followed by massive community transmission and the establishment of local clusters (Figure 6D). Clade 4 (n=107) is characterized by B.1.1.74 sequences and it is more geographically restricted to the Northeast, especially Pernambuco (n=41) and Paraiba (n=23). However, it accounts for occasional introductions to Southern and Southeastern regions (Figure 6E). Clade 5 harbors B.1.1.28 (Clade 5.1) and P.1 (Clade 5.2) sequences. B.1.1.28 sequences are mostly widespread in Northern and Southeastern regions (Figure 6F), and P.1 genomes (n=96) are mostly found in Northern Brazil, especially Amazonas (n=50) and Rondônia (n=15), and Southeast (São Paulo; n=17) (Figure 6H). Clade 6 is also represented by genomes that fall into the B.1.1.28 lineage (n=529). However, it is more widespread in the Southeast (n=436), especially in São Paulo (n=412) and in the Southern region (n=72), especially Rio Grande do Sul (n=65) (Figure 6G). Clade 7 is highly supported by sequences of P.2 lineage (n=204). This clade is also widely distributed throughout Brazilian regions, especially the Southeast (n=83), South (n=63), and North (n=42), even giving rise to new local clusters in the end of 2020 and early-2021 (Figure 6H).

**Figure 6.**
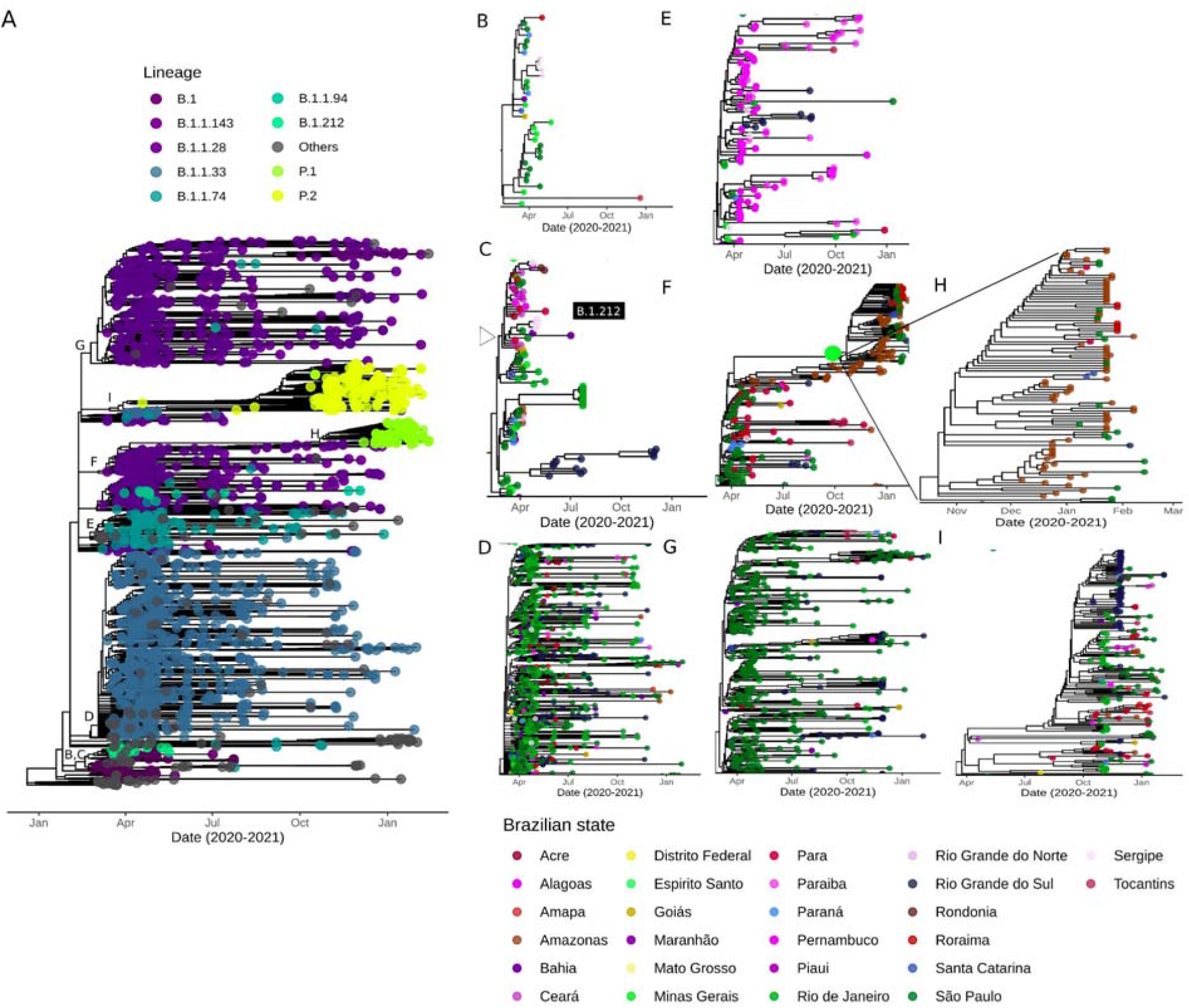
Zoom-in on clades corresponding to the major six Brazilian lineages. (A) Time-resolved ML tree of Brazilian sequences colored by PANGO lineages. Letters around clades are augmented in the respective figures and colored by Brazilian states. (B) Clade 1 is represented by sequences from the B.1 lineage. (C) Clade 2 has sequences from the B.1 and B.1.212 lineages (D) Clade 3 corresponds to the B.1.1.33 lineage. (E) Clade 4 is represented by B.1.1.74 genomes. (F) Clade 5 harbor sequences from B.1.1.28 and P.1. (G) Clade 6 has sequences from the B.1.1.28 lineage. (H) Zoom-in on P.1 sequences from Clade 5. (I) Clade 7 corresponds to the P.2 lineage. From (B) to (I), states belonging to each specific Brazilian region are colored using similar colors (Centre-West: yellow; North: red; Northeast: purple and pink; South: blue; Southeast: green).

### Phylogeographic and phylodynamic patterns of SARS-CoV-2 spread in Brazil

Considering the four major clades mostly represented by Brazilian sequences and with sufficient temporal signal (clades 3 to 6), we reconstructed their spatiotemporal diffusion in the Brazilian territory.

Clade 3 (lineage B.1.1.33) presented a median evolutionary rate of 5.11×10^−4^ (95% Highest Posterior Density (HPD): 4.68×10^−4^ to 5.57×10^−4^). The phylogeographic reconstruction suggests multiple introductions of this clade in different Brazilian regions followed by massive intrastate spread and few transmissions to surrounding states (Figure 7A, Figure S5A, Video S1). It can be illustrated in the MCC tree, which shows five subclades, two mostly represented by sequences from the Southeast, one from the South, one from the Northeast and the other from the North (Figure S5A).

**Figure 7.**
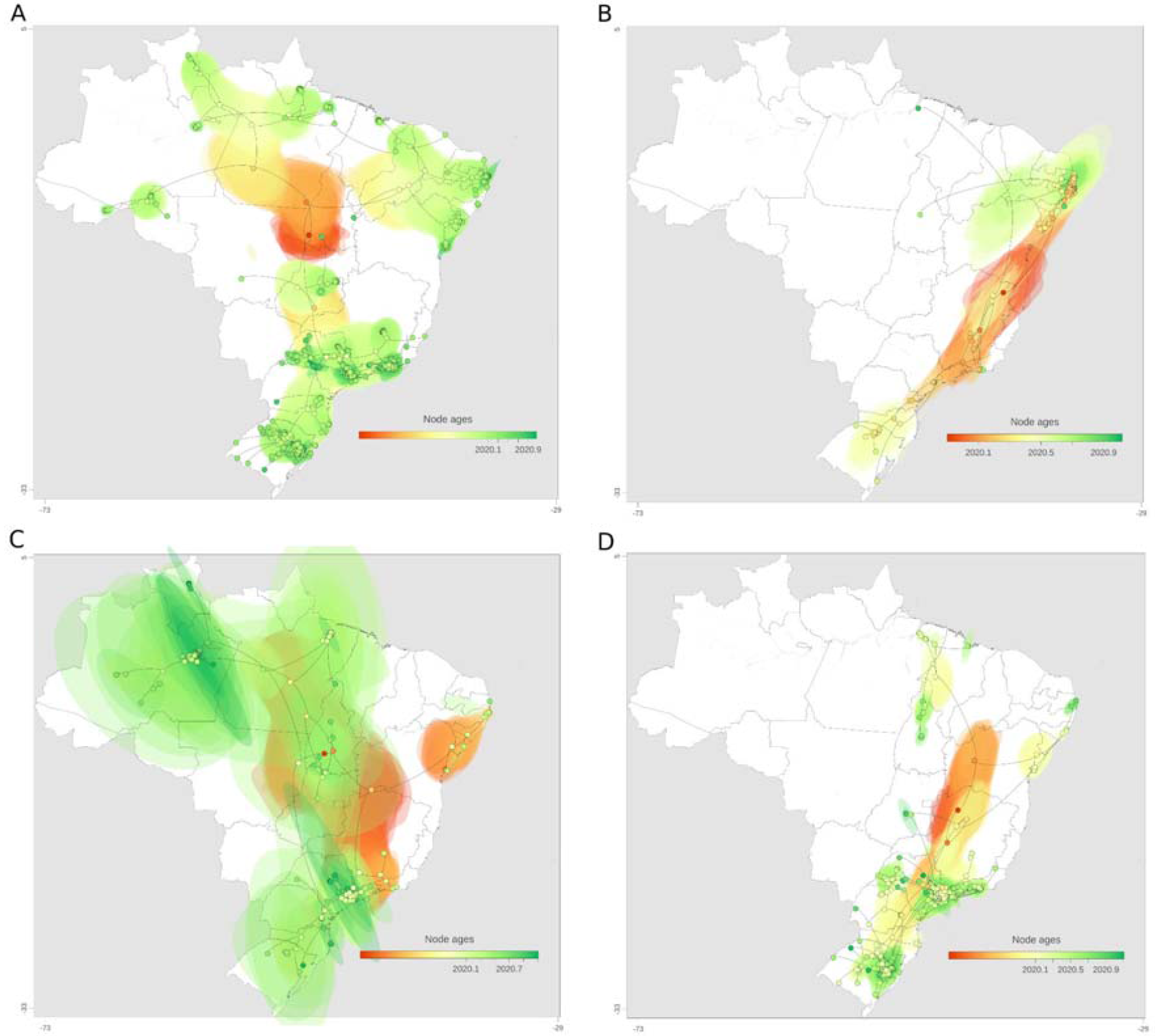
Spatiotemporal reconstruction of the dispersal history of SARS-CoV-2 in Brazil in the first year of the COVID-19 pandemic. Reconstructions of (A) Clade 3, (B) Clade 4, (C) Clade 5, and (D) Clade 6 are represented. MCC trees and 80% HPD regions are based on 1,000 trees subsampled from the posterior distribution of a continuous phylogeographic analysis. Nodes of the MCC tree are coloured according to their time of occurrence. 80% HPD regions were computed for successive time layers and then superimposed using the same colour scale reflecting time.

Clade 4 (lineage B.1.1.74) probably emerged in the Southeast and rapidly spread to Northeast and Southern regions, dispersing progressively from state to state in these regions. A new introduction into the Northeast occurred around September 2020, seeding transmission events to neighboring Northern states (Figure 7B, Figure S5B, Video S2). This clade had a median evolutionary rate of 6.66×10^−4^ (95% HPD: 5.49×10^−4^ to 7.79×10^−4^) and is more geographically restricted to the Brazilian states in the eastern coast, especially states from the Northeast.

Clade 5 (lineages B.1.1.28 and P.1) had a median evolutionary rate of 4.92×10^−4^ (95% HPD: 3.94×10^−4^ to 5.82×10^−4^). This estimate led to an older dating (2018.63, 95% HPD: 2018.03 to 2019.14) of the most recent common ancestor (TMRCA) and instabilities in the tree rooting. Thus, time- or rate-related inferences for this clade should be considered with extreme caution. A hallmark of this clade is its rapid diffusion to all five Brazilian regions (until around May 2020), followed by massive local transmission within the same state or neighboring states in the same region, with higher prevalence in Amazonas (North) and São Paulo (Southeast) (Figure 7C, Figure S5C, Video S3).

Clade 6 (B.1.1.28) had a median evolutionary rate of 6.06×10^−4^ (95% HPD: 5.46×10^−4^ to 6.37×10^−4^). Each of the three subclades within this clade are homogeneously distributed in states from the same region (Figure S5D) and phylogeographic estimates highlighted early introductions in the Southeast, Southern and Northeast regions seeding mostly intra and interstate spread, mainly in the Southeast and Southern Brazil over 2020 (Figure 7D, Video S4).

## Discussion

Viral sequencing is essential to track viral evolution and spread patterns. Despite the initial efforts to obtain a representative genomic dataset of the Brazilian first epidemic wave to better characterize viral introductions and early spread ^17^, the initiatives across different regions have been limited and non-uniform after on. Interestingly, the previously described clades ^17^ have spread and diversified through the country. Clade 1 (after named B.1.1.28) were mostly restricted to the Southeast (São Paulo) and Clade 2 (after named B.1.1.33) were already present in 16 states in this early phase. In this work, we showed the emergence of at least four clades derived from B.1.1.28. The first was widely distributed in Brazilian regions (Clade 5.1) and evolved to P.1 lineage (Clade 5.2). The second (Minor clade 5.3) and third (Clade 6) are most widespread in the Southeast, accounting for a few introductions in other regions. The fourth gave rise to P.2 lineage (Clade 7), which is distributed in all Brazilian regions. B.1.1.33 continues to be composed of a larger clade (Clade 3) with a wide distribution among all Brazilian regions.

The B.1.1.33 lineage was studied in further detail using 190 genomes from 13 Brazilian states, showing its variable abundance in different states (ranging from 2% in Pernambuco to 80% in Rio de Janeiro), and its moderate prevalence in South American countries (5-18%). Surprisingly, this lineage was firstly detected in early-March in other American countries (*e. g*., Argentina, Canada, and USA), and additional analysis suggest that an intermediate lineage (B.1.1.33-like) most probably arose in Europe and was later disseminated to Brazil, where its spread gave origin to lineage B.1.1.33 ^52^ and possibly seeded secondary outbreaks in Argentina and Uruguay ^52,53^.

The states of Pernambuco (Northeast) and Minas Gerais (Southeast) presented more restricted viral dynamics. In Pernambuco, 88% of 101 early sequences were classified as lineage and six local B.1.1 clades were seeded through both national and international traveling ^19^. This finding is consistent with the prevalence of B.1.1.74 lineage found in the Northeast, especially in Pernambuco. In Minas Gerais, 92.5% of the 40 genomes from March 2020 belong to the B lineage (mostly B.1.1) and epidemiological analysis revealed that the distribution of cases and deaths was more spatially uniform, while in other Southeastern states it was more centralized around capital cities ^18^.

Studies from Rio de Janeiro (Southeast) and Rio Grande do Sul (South) identified B.1.1.28 and B.1.1.33 in higher proportion from April to December 2020 ^54–56^. The emergence of a B.1.1.28-derived lineage carrying the E484K mutation (later named P.2) was dated in July 2020, however it began to appear more frequently and almost simultaneously in October 2020 in the Rio de Janeiro state ^54^ and in the small municipality of Esteio, Rio Grande do Sul ^55^, suggesting its wide distribution in the Southern and Southeastern regions of Brazil and uncertainty regarding its origin. This assumption and the frequency increase of B.1.1.28 and derived lineages were corroborated by another study from several municipalities of Rio Grande do Sul, which found that 86% of the sequenced genomes were classified as B.1.1.28 and ∼50% of these belong to the new lineage P.2^56^. Here, we found that P.2 is already distributed in all Brazilian regions up to mid-February 2021.

Recent findings using 250 genomes (March 2020 to January 2021) from Manaus, showed that the first exponential growth phase was driven mostly by the dissemination of B.1.195 which was gradually replaced by B.1.1.28, and the second wave coincides with the emergence of the VOC P.1. This variant probably evolved from a local B.1.1.28 clade in late November and replaced the parental lineage in less than two months. An evolutionary intermediate between B.1.1.28 and P.1 (named P.1-like) was identified, suggesting that the diversity of SARS-CoV-2 variants harboring spike mutations in Manaus could be larger than initially expected and that those variants probably circulated for some time before the emergence and expansion of P.1 ^57^.

Another study following the circulation of P.1 estimated its emergence to November 2020 preceded by a period of faster molecular evolution. Additionally, virus exchanges between Amazonas and the urban metropolises in Southeast Brazil follow patterns in national air travel mobility, since states reporting P.1 until end-February 2021 received around 100,000 air passengers from Manaus in November. Of the 10 new amino acid mutations in the spike protein (L18F, T20N, P26S, D138Y, R190S, K417T, E484K, N501Y, H655Y, T1027I) compared to its immediate ancestor (B.1.1.28), molecular selection analyses found evidence that 9 of these 10 mutations are under diversifying positive selection ^58^.

Detecting mutations that are subjected to positive pressure is of paramount importance in order to predict the SARS-CoV-2 pandemic future. By correlating amino acid replacements with expected structural changes, it is possible to anticipate risk of immune evasion with consequent infection recurrence and or vaccine mismatching. Various specific mutations were encountered in different lineages that are potentially associated with selective advantages. RBD and its hACE-2 interacting core, the Receptor Binding Motif (RBM), is of evident importance, since substitutions in this motif were associated with increased receptor binding forces (*e. g*., N501Y) or immune evasion (*e*.*g*., E484K). The E484K seems to be of particular relevance, as its presence shifts the main interaction residue to this site. Molecular dynamic simulation reveals E484K mutation enhances spike RBD-ACE2 affinity and the combination of E484K, K417N and N501Y mutations (501Y.V2 variant) induces a higher number of conformational changes than N501Y mutant alone, potentially resulting in an escape mutant ^59^. Since this site is not involved in interaction with hACE-2 when the original glutamate is in place, its occurrence has been linked to reinfection, convalescent plasma activity abolishment and decreased post-vaccination neutralizing activity ^59,60^.

Other mutations likely play important roles by allosteric mechanisms and have been positively selected early during the SARS-CoV-2 pandemic. Almost all Brazilian sequences harbor S:D614G, a hallmark of the ancestral B.1 lineage. Although this mutation is outside the RBD, it is speculated that it abolishes the hydrogen bond between the 614 position in S1 and a threonine residue located at S2 from the neighbour protomer. In consequence, RBD would be locked in its activated “up” position, thus increasing viral infectivity ^61^. Therefore, the establishment of this mutation in Brazilian sequences seems to be related to both a founder effect based on the importation of primarily G614 variants to Brazil and an evolutionary advantage in comparison with D614.

The precise forces that drive the appearance of complex mutational signatures characteristic of different lineages over short time periods remain largely unknown. Under specific circumstances, the combination of prolonged viral shedding with high selective pressure could lead to major evolutionary leaps. Critically ill and immunosuppressed patients chronically infected with SARS-CoV-2 and treated with convalescent plasma have been linked to viral breakthroughs caused by mutant viruses ^62^. Whichever phenomena allow for faster viral evolution, they probably have to permit multiple substitutions to occur almost simultaneously, since our phylodynamics analysis shows constant and relatively slow rates of mutation accumulation, except for VOC viruses.

Although the proximal origins of the most important VOCs remain to be determined, some conclusions about their nature could be already drawn. First, the mutations shared between P.1 and B.1.1.351 seem to be associated with a rapid increase in cases even in locations where previous attack rates were thought to be very high ^63^. Lineage P.1, which emerged from the Brazilian state of Amazonas between November and December 2020, has accumulated a high number of non-synonymous mutations and is now dispersed across novel Brazilian regions, representing one of the most frequent lineages up to February 2021 ^22,57^. Second, the fact that the set of mutations shared by P.1, B.1.1.7 and B.1.351 seem to have arisen independently, as we have previously demonstrated with emergence of E484K in others Brazilian lineages (P.2, B.1.1.28 and B.1.1.33), is suggestive of convergent molecular evolution ^58,64^.

Our study shows that the Brazilian territory was affected by at least 59 different lineages during the first year COVID-19 pandemic. This is not completely unexpected, considering the size of the country and its touristic and economical relevance. South Africa, the original source of B.1.351 lineage, similarly had multiple and diverse viral introductions. Of note, a recent genomic study detected 42 different circulating lineages in the country, between the first epidemic wave (March) and mid-September, 2020. Moreover, the three main lineages (B.1.1.54, B.1.1.56 and C.1), which represented the majority of cases in the first wave, were responsible for ∼42% total of the infections by the end of 2020, since B.1.351 had emerged in an explosive fashion in mid-October ^65^. This later lineage is of major concern and South Africa is, up to February 2021, the leader country in COVID-19 related deaths in Africa ^5,66^. In Brazil, the prevalence of lineages B.1.1.28 and its derivatives P.1 and P.2 have been representing progressively more cases of the sequenced genomes by the time of writing. In March 2020, B.1.1.28 was one of the 28 circulating lineages present in the country, with more than 30% of the sequenced genomes. At its side, B.1.1.33 was the identified lineage for approximately 26% of analyzed genomes. Ten months later (January, 2021), the VOC P.1 appeared as the prevalent one among 12 different lineages, reaching more than 45% of the sequenced cases. Next, followed the P.2 lineage (∼27%). At that point, B.1.1.28 and B.1.1.33, which achieved 85.5% of the sequenced genomes together in June, matched for, in January, less than 10% of the cases. Therefore, in both countries, despite potentially different initial founding effects that could have led to diverse lineage dissemination patterns, eventually complex VOCs harboring advantageous mutations were selected for viral spread, indicating an increased evolutionary fitness for these viruses.

Our phylogeographic reconstruction demonstrated the widespread dissemination of multiple SARS-CoV-2 lineages inside Brazil and the major role of intrastate diffusion of the most prevalent lineages. The evolutionary rate estimates for clade-specific MCC trees (especially clades 3, 5, and 6) were significantly smaller than previous findings (8 to 9×10^−4^ subst/site/year) ^67,68^. These differences have contributed to an older dating of the MRCA, since the accumulation of more mutations is expected to occur in a wider timeframe. While these older MRCA datings are probably not realistic in the context of the COVID-19 pandemic, they highlight issues with the data collection process. Potential explanations for this behavior are: related samples having the same age (phylo-temporal clustering), among-lineage rate variation and non-random sampling ^69^.

The dispersion model used is well suited for spread over land, in opposition of long distances traveled by plane, a common practice in the Brazilian territory. The major consequence is that inferences of the nodes locations near to the root of the tree must be analyzed very cautiously. For example, in Figure 7C the green ellipses in the northwest-southeast direction are a consequence of sparse sampling and nodes representing travel events between the North and Southeast. Therefore, the strong uncertainty in these estimates (large ellipses) reflects limitation of both the data and the employed model. Despite its limitations, more local dispersal routes represented here closer to terminal nodes were well captured by the phylogeographic model, which was the main objective of this study since there was a greater contribution of intra- and interstate diffusion after the pandemic was established in the country.

Unfortunately, incomplete and erratic sequencing efforts have limited a better SARS-CoV-2 characterization in Brazil, since genomes are not equally distributed in geographical or temporal scales due to episodic sampling efforts prompted by resource availability. This is reflected by a very small fraction of SARS-CoV-2 cases being sequenced. Unequal temporal distribution implies that some of the conclusions are disproportionately affected by events in heavily (March-May 2020) and poorly (June 2020 onward) sampled periods. Therefore, different lineage distributions could be an artefact of distinct sequencing coverage among states and across different time-frames. Importantly, as Brazil is currently the new epicenter of the pandemic and identified variants of concern and interest, the country initiatives doubled the quantity of genomes deposited in GISAID (5,468 in April 8 vs 2,751 used in this analysis until February 16). Only 420 of these were collected between mid-February and early-April 2021, showing important delays between sample collection and sequencing or sample storage for long periods before sequencing and submission (*e. g*., waiting for reagents and/or research investments). Therefore, a significant amount of sequences from 2020 were deposited lately and were not included in this analysis.

Additionally, the majority of the sequences come from the Southeastern region of Brazil. While this is in fact an economic and travel hub for the country, accounting for >70% of the international passengers arriving in Brazil in the beginning of the pandemic ^70^, inferences regarding this region can be inflated in relation to undersampled regions. However, this is, to the best of our knowledge, the first attempt to characterize sequencing efforts, SARS-CoV-2 mutations, phylogenetics, phylogeography and phylodynamics in the entire Brazil after the study that characterized the first epidemic wave in Brazil using 490 representative genomes ^17^. In the near future, it will be important to describe and track the spread of the P.1 and P.2 lineages, which have already shown to be replacing the other lineages identified in Brazil (https://outbreak.info/situation-reports).

In summary, by systematic analysis of viral genomes distributed across Brazil over time, we were able to confirm the early introductions of multiple lineages, its rapid diversification to constitute new lineages, probable convergent evolution of important mutations (*e. g*., E484K, N501Y), and the emergence of P.1, arguably one of the most potentially concerning lineages identified worldwide up to February 2021. The occurrence of this lineage and the emergence of newer variants could jeopardize the efficacy of vaccines and immunotherapies and may lead the health care system to overload. We concluded that enhanced genomic surveillance is, therefore, of paramount importance and should be extended as soon as possible as a means to better inform policy makers and enable precise evidence-based decisions to fight the COVID-19 pandemic.

## Data Availability

A full table acknowledging the authors and corresponding labs submitting sequencing data used in this study can be found in Supplementary File 1. Additional information used and/or analysed during the current study are available from the corresponding author on reasonable request.

https://www.gisaid.org/

## Acknowledgements

We thank the administrators of the GISAID database and research groups across the world (especially Brazilians) for supporting the rapid and transparent sharing of genomic data during the COVID-19 pandemic. We also thank the Mayor’s Office, Health Department and São Camilo Hospital (Esteio, RS, Brazil), Leonardo Duarte Pascoal and Ana Regina Boll for their work in combating COVID-19 and for supporting the work developed by our research group.

## Declaration of interest statement

The authors declare no competing interests.

## Funding

Scholarships and Fellowships were supplied by the Coordenação de Aperfeiçoamento de Pessoal de Nível Superior – Brasil (CAPES) – Finance Code 001 and Universidade Federal de Ciências da Saúde de Porto Alegre (UFCSPA). The funders had no role in the study design, data generation and analysis, decision to publish or the preparation of the manuscript.

## Author’s contributions

**Vinicius B. Franceschi:** Conceptualization, Methodology, Software, Validation, Formal analysis, Investigation, Data Curation, Writing - Original Draft, Writing - Review & Editing, Visualization. **Patrícia A. G. Ferrareze**: Methodology, Formal analysis, Investigation, Writing - Original Draft, Writing - Review & Editing. **Ricardo A. Zimerman**: Investigation, Writing - Original Draft, Writing - Review & Editing. **Gabriela B. Cybis**: Methodology, Validation, Formal analysis, Investigation, Writing - Review & Editing, Supervision. **Claudia E. Thompson:** Conceptualization, Methodology, Formal analysis, Investigation, Resources, Writing - Original Draft, Writing - Review & Editing, Supervision, Project administration.

## Supplementary files

**Supplementary File 1**. GISAID acknowledgement table of worldwide SARS-CoV-2 genomes used in this study.

**Video S1**. Animation of the phylogeographic reconstruction of Clade 3. Node polygons and lines represent posterior probabilities (orange to red scale). Colors on the map represent the five Brazilian regions (South, Southeast, Centre-West, Northeast and North).

**Video S2**. Animation of the phylogeographic reconstruction of Clade 4. Node polygons and lines represent posterior probabilities (orange to red scale). Colors on the map represent the five Brazilian regions (South, Southeast, Centre-West, Northeast and North).

**Video S3**. Animation of the phylogeographic reconstruction of Clade 5. Node polygons and lines represent posterior probabilities (orange to red scale). Colors on the map represent the five Brazilian regions (South, Southeast, Centre-West, Northeast and North).

**Video S4**. Animation of the phylogeographic reconstruction of Clade 6. Node polygons and lines represent posterior probabilities (orange to red scale). Colors on the map represent the five Brazilian regions (South, Southeast, Centre-West, Northeast and North).

## Notes

### Competing Interest Statement

The authors have declared no competing interest.

### Funding Statement

Scholarships and Fellowships were supplied by the Coordenacao de Aperfeicoamento de Pessoal de Nivel Superior - Brasil (CAPES) - Finance Code 001 and Universidade Federal de Ciencias da Saude de Porto Alegre. The funders had no role in the study design, data generation and analysis, decision to publish or the preparation of the manuscript.

### Author Declarations

We used publicly available genomes from the GISAID initiative following its guidelines.

### Summary of Updates

We have added Bayesian phylogeographic and phylodynamic analyses and improved the text and figures.

