## Supplementary Material for "Mutation hotspots, geographical and temporal distribution of SARS-CoV-2 lineages in Brazil, February 2020-2021: insights and limitations from uneven sequencing efforts"

### **Table of Contents**

**Table S1.** Comparison of genomes sampled from Brazil and confirmed cases per month from March 2020 to mid-February 2021.	2

**Table S2.** Comparison of genomes sampled from each Brazilian state and confirmed cases from March 2020 to mid-February 2021. Data are ordered from the higher sequencing rate (genomes per case) to lower.	3

**Table S3.** Number of genomes and lineages sampled from each Brazilian state from March 2020 to mid-February 2021. Data are ordered from the higher number of genomes to lower.	4

**Table S4.** First and last detection of SARS-CoV-2 lineages represented by > 5 Brazilian genomes, including information on state of detection and minimum time of spread according to available data.	5

**Figure S1.** Distribution of the 10 most prevalent SARS-CoV-2 lineages across all Brazilian states considering the absolute number of sequences per state.	7

**Figure S2.** Time between first and last detection of the 18 lineages observed in more than 5 Brazilian genomes.	8

**Figure S3.** Time-resolved Maximum likelihood phylogenetics tree dropping sequences from other countries and highlighting the most frequent Brazilian **lineages.**	**9**

**Figure S4.** Zoom-in on clades corresponding to the two minor clades from B.1.1.74 and B.1.1.28 Brazilian lineages not represented in Figure 6. (A) Time-resolved ML tree of Brazilian sequences colored by PANGO lineages. Letters around clades are augmented in respective figures. (B) Clade 4.2 is represented by 27 sequences (B.1.1.74 lineage). (C) Clade 5.3 harbor 162 sequences from B.1.1.28 and B.1.1.94.	10

**Figure S5.** Maximum clade credibility trees of major Brazilian clades with sufficient temporal signal. (A) Clade 3 (lineage B.1.1.33), (B) Clade 4 (B.1.1.74), (C) Clade 5 (B.1.1.28 and P.1) and (D) Clade 6 (B.1.1.28).	11

#### **Table S1**. Comparison of genomes sampled from Brazil and confirmed cases per month from March 2020 to mid-February 2021.

| **Year-month** | **Cases** | **Genomes** | **Genomes per case (%)** |
| --- | --- | --- | --- |
| 2020-03 | 5822 | 503 | 8.6396 |
| 2020-04 | 81302 | 942 | 1.1586 |
| 2020-05 | 429011 | 250 | 0.0583 |
| 2020-06 | 896532 | 131 | 0.0146 |
| 2020-07 | 1257782 | 170 | 0.0135 |
| 2020-08 | 1244378 | 94 | 0.0076 |
| 2020-09 | 902536 | 39 | 0.0043 |
| 2020-10 | 720398 | 85 | 0.0118 |
| 2020-11 | 804202 | 226 | 0.0281 |
| 2020-12 | 1339067 | 130 | 0.0097 |
| 2021-01 | 1527489 | 137 | 0.0090 |
| 2021-02 | 658765 | 19 | 0.0029 |

* February 2020 was excluded due to more number of genomes (6) than confirmed cases (2). February 2021 data until day 16 were included.

#### **Table S2**. Comparison of genomes sampled from each Brazilian state and confirmed cases from March 2020 to mid-February 2021. Data are ordered from the higher sequencing rate (genomes per case) to lower.

| **Region** | **State** | **Abbrev. State** | **Genomes** | **Cases** | **Genomes per case (%)** |
| --- | --- | --- | --- | --- | --- |
| Southeast | Rio de Janeiro | RJ | 496 | 555541 | 0.0893 |
| Southeast | São Paulo | SP | 1108 | 1915914 | 0.0578 |
| South | Rio Grande do Sul | RS | 264 | 586315 | 0.0450 |
| North | Amazonas | AM | 121 | 295879 | 0.0409 |
| Northeast | Pernambuco | PE | 114 | 281331 | 0.0405 |
| Northeast | Paraíba | PB | 81 | 205579 | 0.0394 |
| North | Amapá | AP | 27 | 80663 | 0.0335 |
| North | Pará | PA | 101 | 348448 | 0.0290 |
| North | Acre | AC | 10 | 53455 | 0.0187 |
| Northeast | Sergipe | SE | 26 | 145677 | 0.0178 |
| North | Roraima | RR | 13 | 77531 | 0.0168 |
| North | Rondônia | RO | 23 | 137384 | 0.0167 |
| North | Tocantins | TO | 15 | 107240 | 0.0140 |
| Southeast | Minas Gerais | MG | 98 | 808693 | 0.0121 |
| Northeast | Maranhão | MA | 25 | 213087 | 0.0117 |
| Northeast | Ceará | CE | 41 | 396495 | 0.0103 |
| Northeast | Bahia | BA | 54 | 631645 | 0.0085 |
| South | Paraná | PR | 35 | 589494 | 0.0059 |
| Northeast | Rio Grande do Norte | RN | 9 | 156939 | 0.0057 |
| Northeast | Alagoas | AL | 7 | 125044 | 0.0056 |
| Centre-West | Goiás | GO | 15 | 372573 | 0.0040 |
| Centre-West | Distrito Federal | DF | 11 | 285576 | 0.0039 |
| South | Santa Catarina | SC | 20 | 615441 | 0.0032 |
| Centre-West | Mato Grosso | MT | 6 | 233091 | 0.0026 |
| Northeast | Piauí | PI | 2 | 166098 | 0.0012 |
| Southeast | Espírito Santo | ES | 2 | 311236 | 0.0006 |
| Centre-West | Mato Grosso do Sul | MS | 1 | 170917 | 0.0006 |

#### **Table S3**. Number of genomes and lineages sampled from each Brazilian state from March 2020 to mid-February 2021. Data are ordered from the higher number of genomes to lower.

| **Region** | **State** | **Abbrev. State** | **Genomes** | **Lineages** | **All lineages** |
| --- | --- | --- | --- | --- | --- |
| Southeast | São Paulo | SP | 1,108 | 33 | B.1.1, B.1, B.1.1.74, B.1.1.33, B.1.1.28, B.1.1.143, B.1.212, B.1.1.94, B, B.1.1.244, B.1.1.314, B.1.146, B.1.1.161, B.1.1.44, B.1.1.304, N.4, B.1.1.208, B.1.1.7, B.1.195, P.2, B.1.1.10, N.1, P.1, B.1.177, B.1.1.162, B.3, B.1.258, B.1.1.307, B.1.1.288, B.1.1.222, B.23, B.40 |
| Southeast | Rio de Janeiro | RJ | 496 | 17 | B.1, B.1.1.74, B.1.1.33, B.39, B.1.1.143, B.1.1.28, A.2, B.1.1.277, P.2, B.1.1.296, B.1.1.314, N.1, N.4, B.1.1.279, B.1.1.161, B.1.1.54, B.1.1.10 |
| South | Rio Grande do Sul | RS | 264 | 16 | B.1, B.1.1.33, B.1.1.28, P.2, B.1.91, B.1.1.143, B.1.1.279, B.1.1.314, B.1.1.74, B.1.1.107, P.1, B.1.1.161, A.5, B, B.1.1.10, B.1.1.94 |
| North | Amazonas | AM | 121 | 10 | B.1.195, B.1.1.33, B.1.1.28, P.1, P.2, B.1.1, B.1.1.74, B.1.1.250, B.1.1.288, A.2 |
| Northeast | Pernambuco | PE | 114 | 10 | B.1.1.74, B.1.1.33, B.1.1.28, B.1, B.1.1.117, B, B.1.212, B.1.1.298, B.1.1.214, B.1.1.107 |
| North | Pará | PA | 101 | 12 | B.1.1.28, B.1.1.33, B.1.1.1, B.1.1.74, P.1, B.1.212, P.2, B.39, B.1.225, B.1, B.1.1.4, B.1.1.143 |
| Southeast | Minas Gerais | MG | 98 | 14 | B.1, B.40, B, B.1.1.74, B.1.1.33, B.1.314, B.1.1.314, B.1.1.161, B.1.1.10, B.1.1.1, B.1.1.28, B.1.212, B.1.1.71, B.1.258 |
| Northeast | Paraíba | PB | 81 | 9 | B.1.212, B.1.1.74, B.1.1.33, P.2, B.1.1.145, N.4, B.1.1.28, B.1.1.141, B.1.1.291 |
| Northeast | Bahia | BA | 54 | 10 | B.1, B.1.1.33, P.2, B.1.1.314, B.1.1.161, A.1, B.1.1.28, B.1.1.162, B.3, B.1.1.94 |
| Northeast | Ceará | CE | 41 | 6 | B.1.212, B.1.1.33, B.1, B.1.1.28, P.2, B.1.1.10 |
| South | Paraná | PR | 35 | 7 | B.1.1.74, B.1.1.28, B.1.1.33, B.1.195, B.1, P.2, B.1.1.54 |
| North | Amapá | AP | 27 | 4 | B.1.1.33, N.2, P.2, B.1.160 |
| Northeast | Sergipe | SE | 26 | 5 | B.1.1.74, B.1, B.1.1.33, B.1.1.28, B.1.212 |
| Northeast | Maranhão | MA | 25 | 1 | B.1.1.33 |
| North | Rondônia | RO | 23 | 3 | B.1.1.33, B.1.212, P.1 |
| South | Santa Catarina | SC | 20 | 6 | B.6, B.1, B.1.1.33, B.1.1.28, B.1.1.74, P.1 |
| North | Tocantins | TO | 15 | 5 | B.1.1.74, B.1.1.28, B.1.1.33, B.1.1.4, P.2 |
| Centre-West | Goiás | GO | 15 | 7 | B.1, B.40, B.1.22, B.1.1.33, B.1.1.7, B.1.1.28, P.2 |
| North | Roraima | RR | 13 | 3 | B.1.1.33, P.1, B.1 |
| Centre-West | Distrito Federal | DF | 11 | 5 | B, B.1.1.314, B.1.1.33, P.2, B.1.1.7 |
| North | Acre | AC | 10 | 2 | B.1.1.33, B.1.212 |
| Northeast | Rio Grande do Norte | RN | 9 | 3 | B.1.195, B.1.1.33, B.1 |
| Northeast | Alagoas | AL | 7 | 4 | B.1.1.28, P.2, B.1.1.33, B.1.1.1 |
| Centre-West | Mato Grosso | MT | 6 | 3 | B.4, B.1.1.33, B.1.1.28 |
| Southeast | Espírito Santo | ES | 2 | 2 | B.40, B.1.1.33 |
| Northeast | Piauí | PI | 2 | 1 | B.1.1.33 |
| Centre-West | Mato Grosso do Sul | MS | 1 | 1 | B.1.1.33 |

#### **Table S4**. First and last detection of SARS-CoV-2 lineages represented by > 5 Brazilian genomes, including information on state of detection and minimum time of spread according to available data.

| **Lineage** | **Genomes** | **First time detected** | **State** | **Last time detected** | **State** | **Minimum time of spread** |
| --- | --- | --- | --- | --- | --- | --- |
| B | 12 | 2020-02-28 | São Paulo | 2020-04-09 | Pernambuco | 41 |
| B.1 | 167 | 2020-02-25 | São Paulo | 2020-07-28 | Rio de Janeiro | 154 |
| B.1.1.10 | 6 | 2020-03-18 | Minas Gerais | 2021-02-09 | São Paulo | 328 |
| B.1.1.143 | 60 | 2020-03-19 | São Paulo | 2021-02-01 | Rio Grande do Sul | 319 |
| B.1.1.161 | 10 | 2020-03-18 | Minas Gerais | 2021-01-08 | São Paulo | 296 |
| B.1.1.162 | 7 | 2020-02-28 | Bahia | 2020-05-30 | Bahia | 92 |
| B.1.1.28 | 882 | 2020-03-05 | São Paulo | 2021-02-09 | São Paulo | 341 |
| B.1.1.314 | 28 | 2020-03-13 | Distrito Federal | 2020-08-24 | Rio Grande do Sul | 164 |
| B.1.1.33 | 866 | 2020-03-09 | São Paulo | 2021-01-27 | São Paulo | 324 |
| B.1.1.7 | 19 | 2020-12-21 | São Paulo | 2021-01-22 | São Paulo | 32 |
| B.1.1.74 | 194 | 2020-03-04 | São Paulo | 2020-12-25 | Pará | 296 |
| B.1.1.94 | 24 | 2020-03-25 | São Paulo | 2020-11-30 | São Paulo | 250 |
| B.1.195 | 12 | 2020-03-17 | Paraná | 2020-04-17 | Rio Grande do Norte | 31 |
| B.1.212 | 25 | 2020-03-14 | Ceará | 2020-05-13 | Pará | 60 |
| B.1.91 | 11 | 2020-05-20 | Rio Grande do Sul | 2020-11-30 | Rio Grande do Sul | 194 |
| B.39 | 9 | 2020-03-10 | Rio de Janeiro | 2020-05-18 | Rio de Janeiro | 69 |
| P.1 | 109 | 2020-12-04 | Amazonas | 2021-02-11 | São Paulo | 69 |
| P.2 | 202 | 2020-04-15 | Ceará | 2021-02-02 | Rio Grande do Sul | 293 |

#### **Figure S1**. Distribution of the 10 most prevalent SARS-CoV-2 lineages across all Brazilian states considering the absolute number of sequences per state.

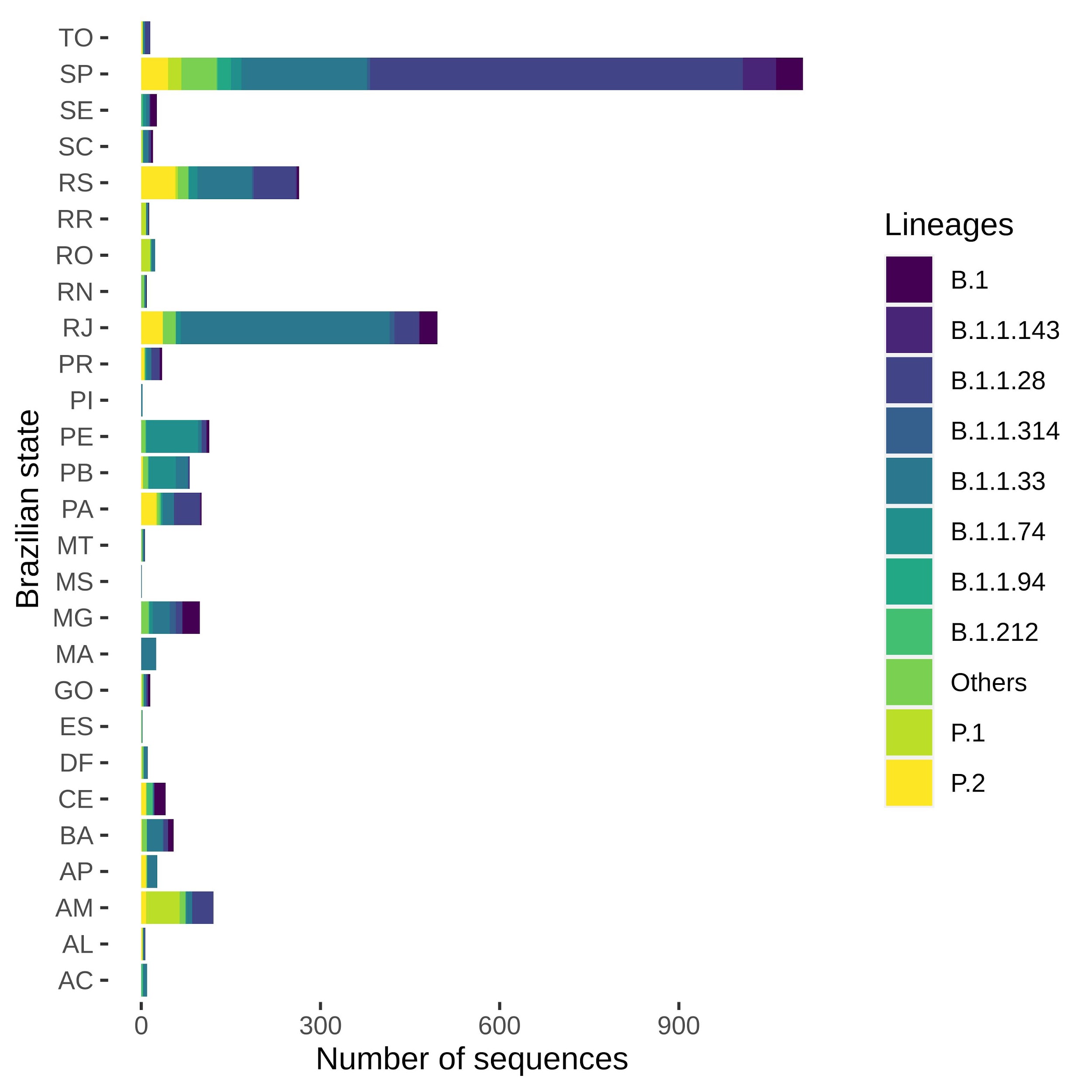

#### **Figure S2**. Time between first and last detection of the 18 lineages observed in more than 5 Brazilian genomes.

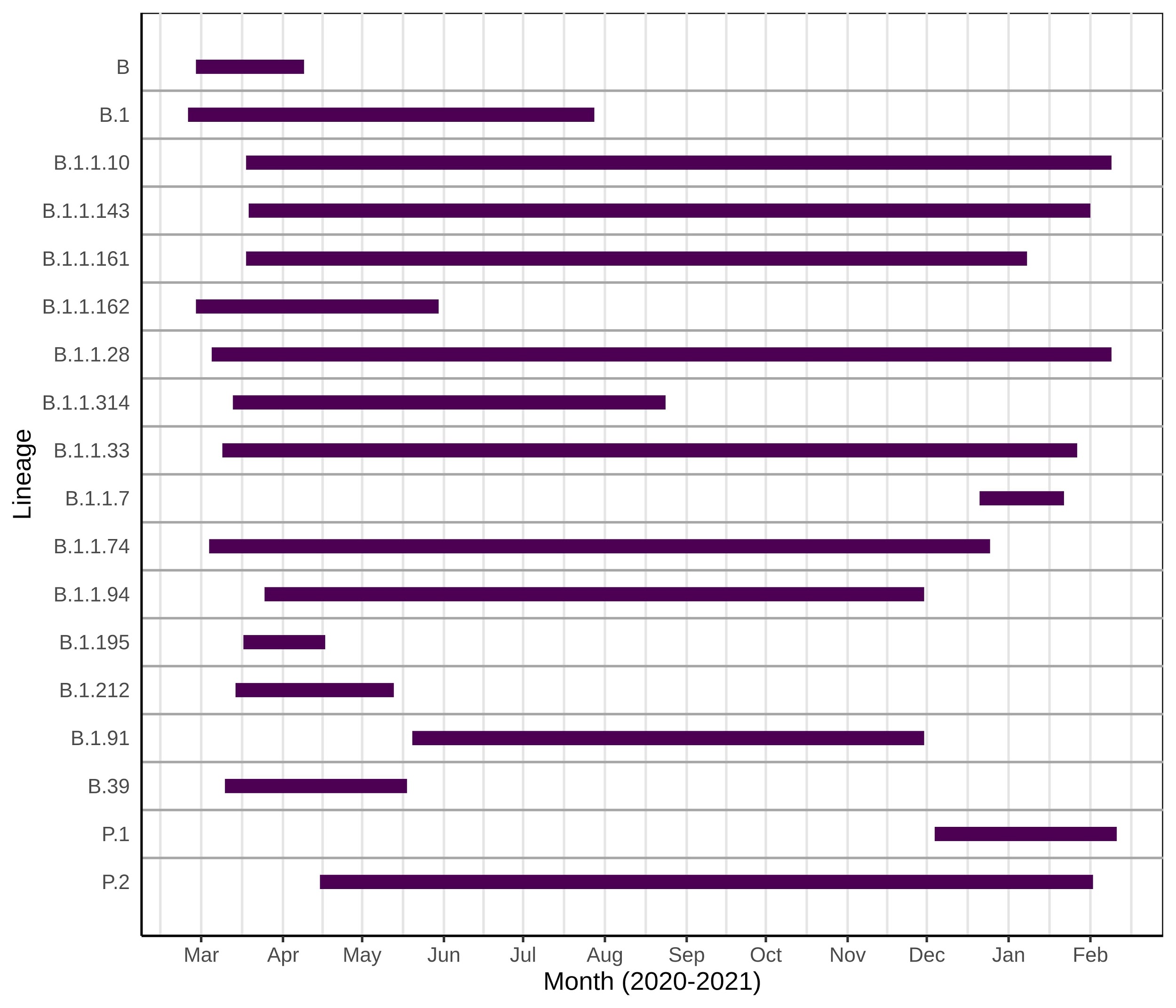

#### **Figure S3.** Time-resolved Maximum likelihood phylogenetics tree dropping sequences from other countries and highlighting the most frequent Brazilian lineages.

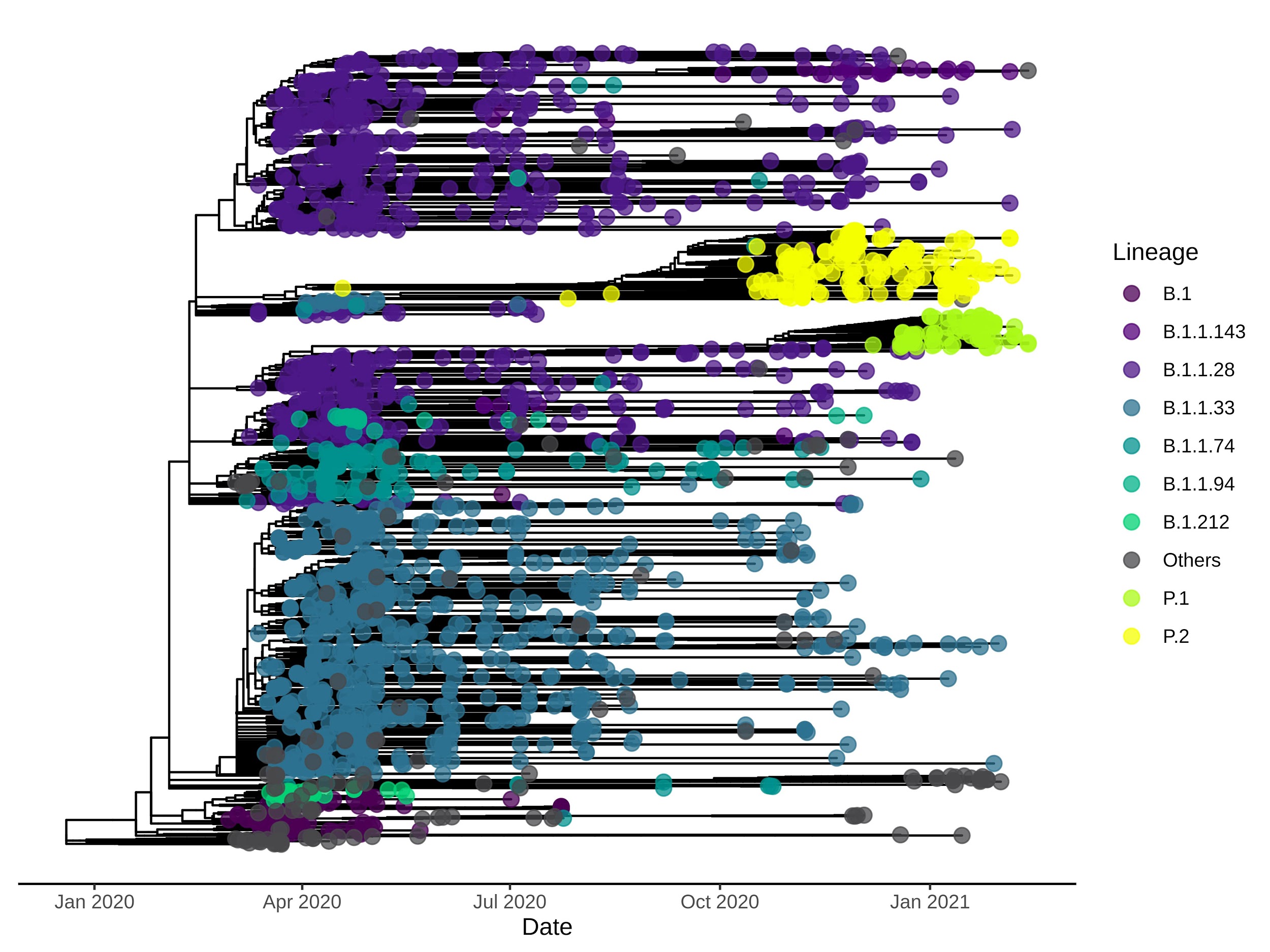

#### **Figure S4.** Zoom-in on clades corresponding to the two minor clades from B.1.1.74 and B.1.1.28 Brazilian lineages not represented in Figure 6. (A) Time-resolved ML tree of Brazilian sequences colored by PANGO lineages. Letters around clades are augmented in respective figures. (B) Clade 4.2 is represented by 27 sequences (B.1.1.74 lineage). (C) Clade 5.3 harbor 162 sequences from B.1.1.28 and B.1.1.94.

#### In (B) and (C), states belonging to each specific Brazilian region are colored using similar colors (Centre-West: yellow; North: red; Northeast: purple and pink; South: blue; Southeast: green).

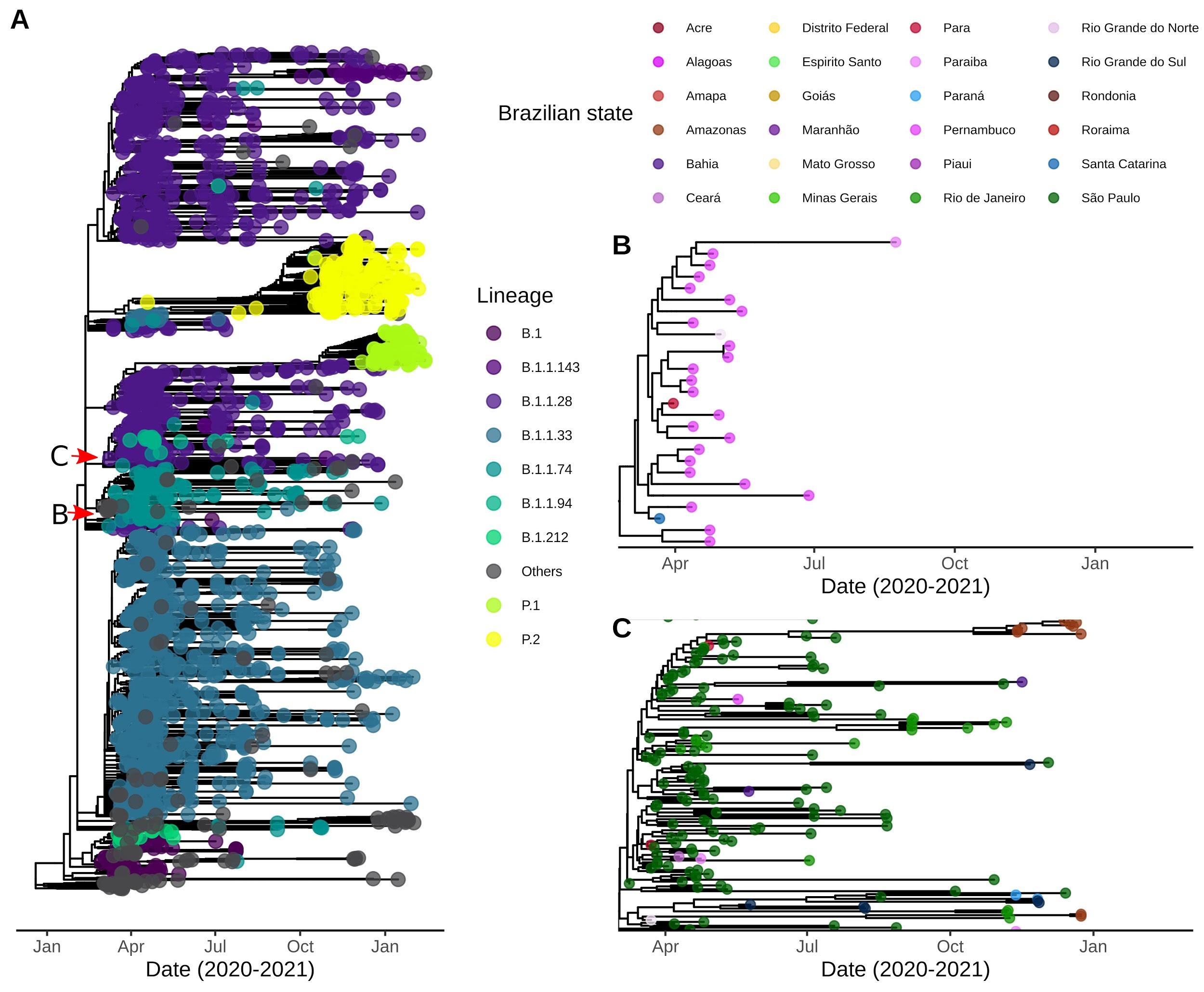

#### **Figure S5.** Maximum clade credibility trees of major Brazilian clades with sufficient temporal signal. (A) Clade 3 (lineage B.1.1.33), (B) Clade 4 (B.1.1.74), (C) Clade 5 (B.1.1.28 and P.1) and (D) Clade 6 (B.1.1.28).

#### In (A), (B), (C), and (D), states belonging to each specific Brazilian region are colored using similar colors (Centre-West: yellow; North: red; Northeast: purple and pink; South: blue; Southeast: green).

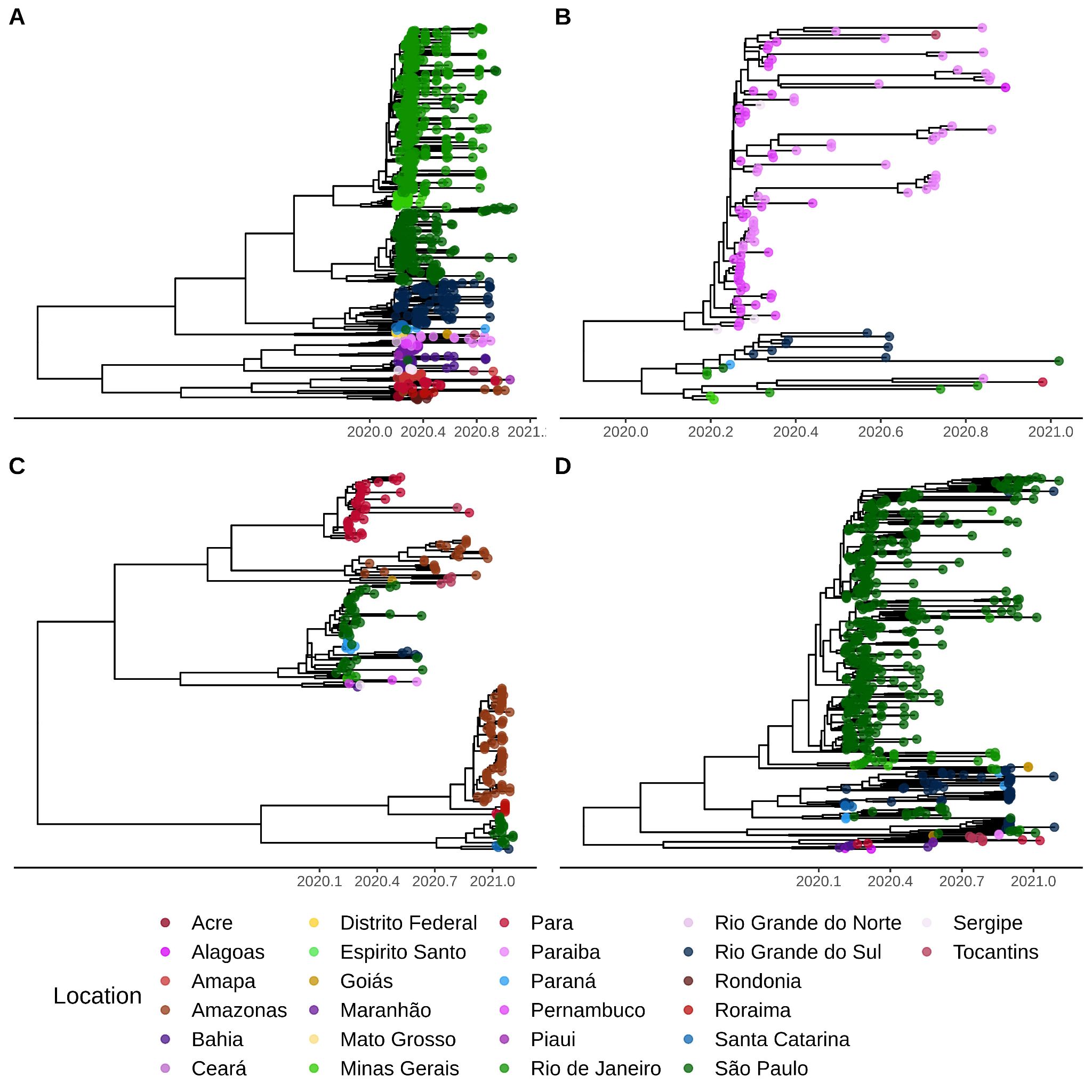
